# Tomato-Soy Juice Reduces Inflammation and Modulates Urinary Metabolome in Adults with Obesity

**DOI:** 10.1101/2025.09.11.25334465

**Authors:** Maria J. Sholola, Jenna Miller, Emma A. Bilbrey, Janet A. Novotny, David M. Francis, Thomas A. Mace, Jessica L. Cooperstone

**Affiliations:** Department of Food Science and Technology, The Ohio State University, Columbus, OH; Department of Horticulture and Crop Science, The Ohio State University, Columbus, OH; Beltsville Human Nutrition Research Center, Agricultural Research Service, U.S. Department of Agriculture, Beltsville, MD; Division of Gastroenterology, Hepatology, and Nutrition, The Ohio State University Wexner Medical Center, Columbus, OH; Department of Horticulture and Crop Science, The Ohio State University, Wooster, OH

## Abstract

**Scope:** Chronic, low-grade inflammation is a hallmark of many noncommunicable diseases, including obesity. Diets enriched with tomatoes and soy have been associated with better health outcomes in inflammation-related illnesses, with lycopene and isoflavones considered key bioactive components, respectively. On the basis that whole food combinations may exert greater effects than isolated phytochemicals, we examine the anti-inflammatory and metabolic effects of tomato-soy juice compared to a low carotenoid tomato juice control in obesity.

**Methods and results:** In a randomized, crossover trial, 12 healthy adults with obesity were provided either tomato-soy juice (54 mg lycopene/d, 189.9 mg isoflavones/d) or a low carotenoid tomato juice (no isoflavones) daily for 4 weeks, then crossed over to the other treatment following a washout period. Plasma carotenoids, cytokines, and the urine metabolome were measured pre- and post-interventions. Plasma lycopene significantly increased by 2.48-fold after tomato-soy intake. IL-5, IL-12p70, and GM-CSF significantly decreased (*P* < 0.05), and TNF-α trended downward (*P* = 0.052) following tomato-soy. Soy isoflavones and their metabolites primarily distinguished post-tomato-soy urine profiles. Both interventions induced some shared metabolomic changes in the urine, indicating tomato-driven effects independent of lycopene.

**Conclusion:** Tomato-soy intake reduced some pro-inflammatory cytokines and altered the urine metabolomic profile in adults with obesity, supporting future studies using this functional food product for other inflammation-related conditions.

## 1 Introduction

While inflammation is an essential mechanism in the body for immune defense, its persistent activation can impair physiological functions by promoting oxidative stress and tissue damage. In 2018, at least half of American adults were diagnosed with one or more chronic illness linked to inflammation (Boersma, 2020). Obesity, for example, affects ∼42% of adults over 20 years old in the U.S (Stierman et al., 2021). Not only is obesity a major public health concern on its own, it is also a risk factor for other burdensome chronic diseases including cardiometabolic disease (Valenzuela et al., 2023) and cancer (Wolin et al., 2010). Low-grade inflammation is also common hallmark in other noncommunicable diseases (Hotamisligil, 2017; Plutzky, 2001) and is characterized by high, but subclinical, levels of cytokines such as IL-6, tumor necrosis factor (TNF)-α, IL-1β, and C-reactive protein (CRP) (Dinarello, 2006; Emanuela et al., 2012; Glas et al., 2004; Ridker et al., 2000; Rodríguez-Hernández et al., 2013).

Diet is recognized as a modifiable factor which has consistently shown to be correlated with reduced prevalence of chronic diseases (McCullough et al., 2002). Aside from essential nutrients such as vitamins and fiber, dietary plants are a major source of non-nutrient phytochemicals— secondary metabolites that function to protect and benefit the host plant which could also promote human health (Krzyzanowska et al., 2010). For example, in plants, carotenoids like lycopene (the red pigment in tomatoes) play a protective role by quenching of reactive oxygen species during photosynthesis (Cunningham & Gantt, 1998; Frank & Cogdell, 1996), and polyphenols such as flavonoids are produced in response to environmental stressors, such as UV radiation, pests and pathogens (Naikoo et al., 2019). Observational studies suggest that phytochemical-rich diets may prevent obesity, cardiometabolic disease, and cancer incidence (Baena Ruiz & Salinas Hernández, 2016; Ghoreishy et al., 2021; Im et al., 2020; Mehranfar et al., 2022; Vasmehjani et al., 2021). Phytochemicals like carotenoids and flavonoids are thought to offer pharmacological properties, acting on inflammatory pathways and other etiological factors associated with chronic disease. In obesity animal models, lycopene supplementation has been shown to inhibit key pro-inflammatory markers from the nuclear factor κB (NF-κB) pathway in adipose and liver tissues by modulating IκB phosphorylation (Fenni et al., 2017; Gouranton et al., 2011). Dietary flavonoids such as those from tomato (e.g., naringenin) and those from soy (e.g., genistein) have also been associated with anti-inflammatory activities by inhibiting signaling of transcription factors and kinases such as NF-κB and extracellular signal-regulated kinase (ERK) in adipocytes (Shen et al., 2019; Yoshida et al., 2010).

Research has increasingly emphasized the role of overall dietary patterns in shaping health outcomes, given that people consume whole foods and not isolated, individual compounds (R. H. Liu, 2003, 2023). Utilizing combinations of foods and phytochemicals for interventions not only reflect realistic eating behaviors, but also capture the interactions between molecules that might influence their biological activity. In this context, we focus on tomatoes and soy, which are both associated with health protective effects, especially in prostate cancer. In 1995, a landmark observational study reported that men who consume at least 10 servings per week of tomato products had a 35% lower risk of advanced prostate cancer in comparison to those who consumed less than 1.5 servings (Giovannucci et al., 1995). Around the same time, prostate cancer rates were reported to be much higher in America than in Japan and China, and soy-rich diets are thought to be one of the dietary factors for this observation (Watanabe et al., 2000; Yatani et al., 1989; YU et al., 1991). These epidemiological studies led to a series of investigations supplementing tomato and soy to target prostate cancer. In a rat prostate carcinoma model, tomato and soy polyphenols (genistein, daidzein, biochanin A, quercetin, rutin, and kaempferol administered individually) were found to inhibit growth factor signaling cascades essential for prostate cancer progression (S. Wang et al., 2003), indicating potential mechanisms for tomato or soy consumption in prostate cancer. In a prostate cancer mouse model, Zuniga et al. (2013) interestingly found that mice fed diets enriched with both tomato and soy germ were more protected against prostate cancer incidence than mice fed soy or tomato powder alone. This suggests additive effects of tomato and soy when combined. To test this synergy in humans, Ohio State researchers developed a novel functional food product, tomato-soy juice (Tiziani & Vodovotz, 2005). Its safety and compliance was later established in healthy men and women (Bohn et al., 2013). Subsequent work by Grainger et al. (2019) demonstrated excellent compliance and plasma, prostatic, and urinary concentrations of carotenoids and isoflavones in men with prostate cancer after consuming varying doses of tomato-soy juice (0, 1, or 2 cans/day). In the same study, there was a dose-dependent trend towards reduced prostate-specific antigen (PSA) levels with increasing tomato-soy juice intake after the short intervention of only 3 weeks (Grainger et al., 2019). Several years before, the same author reported PSA doubling time was prolonged in prostate cancer patients consuming a combination of tomato products and soy protein (Grainger et al., 2008).

While tomato-soy interventions were first applied to prostate cancer, additional promising evidence suggests tomatoes and soy can impact broader inflammatory and metabolic pathways relevant to obesity and other chronic diseases when consumed separately (J. Ahn-Jarvis et al., 2020; Fenni et al., 2017; Ghavipour et al., 2013; Tsuhako et al., 2020) and combined (Mukherjee et al., 2020). Given the overlap of biological pathways shared by cancer, obesity, and inflammation, especially those involving cytokine signaling and metabolism, there is growing interest in exploring tomato-soy interventions in other inflammation-related contexts.

In this study, we investigate the anti-inflammatory and metabolic effects of a high lycopene tomato-soy juice intervention compared to a low-carotenoid tomato juice control in individuals with obesity. The control intervention made from a low-carotenoid tomato variety was intended to serve as a matched matrix, helping to isolate the effects of lycopene and soy while controlling for non-carotenoid tomato phytochemicals. To capture systemic changes in response to intervention, we measured inflammatory cytokines in blood and profiled urinary metabolites using an untargeted metabolomics approach. Analysis of urine offers important insight as it reflects the excretion of both diet-derived and endogenous metabolites in response to each intervention, enabling the sensitive detection of dietary intake to identify metabolic changes and specific phytochemical metabolites linked to tomato-soy consumption. By investigating both inflammatory markers and urinary metabolomic profiles, we aim to uncover molecular signatures and mechanisms that occur following tomato-soy consumption.

## 2 Experimental Section

### 2.1 Subjects and experimental design

A total of 13 subjects began this study, while 1 dropped due to noncompliance during the intermediary washout period (**Figure 1**).

**Figure 1.**
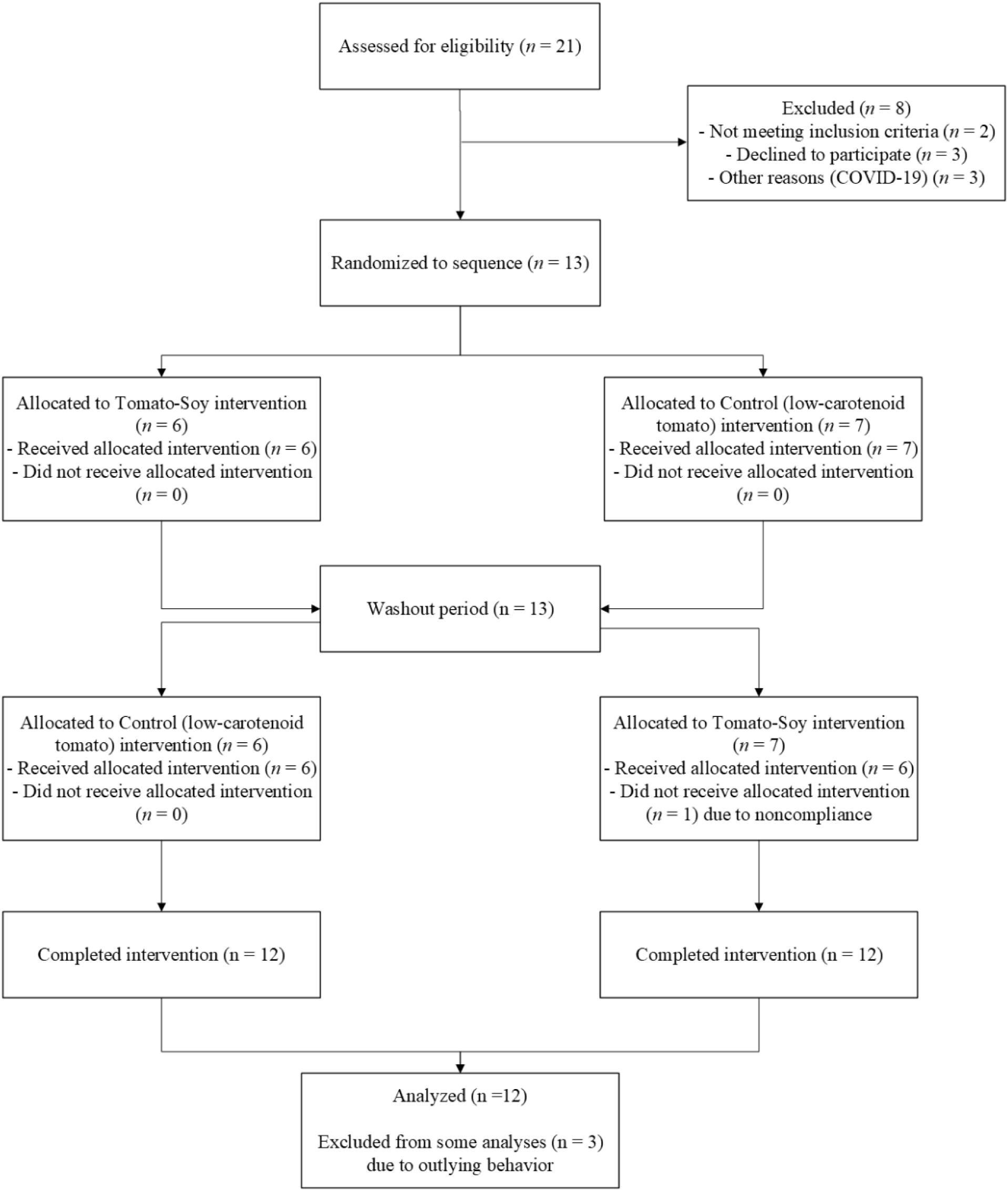
Subject selection CONSORT diagram

Twelve male and female healthy participants with obesity (BMI 30-45 kg/m^2^) in the Beltsville, MD area completed a randomized, crossover trial completed at the Beltsville Human Nutrition Research Center (BHNRC), part of the United States Department of Agriculture, Agricultural Research Service (USDA-ARS). This 14-week study was carried out between March – May 2019. Subjects, aged 30-60 years, were required to have plasma total cholesterol levels ≤ 250 mg/dL and triglyceride levels ≤ 250 mg/dL. Exclusion criteria included smoking, the use of daily prescription anti-inflammatory drugs, antibiotics within 3 months prior to the study, use of carotenoid/isoflavone/metabolism-altering supplements within 1 month of the study, presence of autoimmune or metabolic disorders, presence of any gastrointestinal or malabsorption conditions, indication of liver or kidney disorders, and tomato or soy allergies. Baseline characteristics for participants can be found in **Table 1**.

**Table 1.** Participant characteristics at initial screening visit (mean ± SD). There were no significant differences in subjects’ characteristics by sex.

| Sex | Participants | Age | BMI | Plasma total cholesterol | Plasma triglycerides |
| --- | --- | --- | --- | --- | --- |
|  | *n* | *y* | *kg/m2* | *mg/dL* | *mg/dL* |
| F | 5 | 59.2 ± 11.7 | 35.0 ± 5.8 | 210.3 ± 33.4 | 116.2 ± 43.2 |
| M | 7 | 52.3 ± 12.2 | 32.0 ± 2.4 | 187.3 ± 52.5 | 115.7 ± 74.1 |

To test the effects of both lycopene and isoflavones on inflammation and the metabolome, subjects were instructed to consume diets with low levels of these phytochemicals for 2 weeks prior to the intervention. Counseling and a list of foods to avoid was provided to achieve a low lycopene/isoflavone diet. Following the 2-week washout period, subjects were randomized to receive 360 mL (two 180 mL cans) of either a soy-enriched, high lycopene tomato (tomato-soy) juice or a low carotenoid, yellow tomato juice (control) every day for 4 weeks.

Followed by a 4-week washout period (low lycopene and isoflavone diet), participants began their second 4-week intervention consuming the other juice daily (Figure 2). Fasted blood samples collected from participants at weeks 0, 2, 6, 10, and 14 were centrifuged so that plasma could be aliquoted into cryovials for storage at −80 °C. At weeks 2, 6, 10, and 14, 24-hour urine samples were collected from the subjects into 4 L plastic jugs containing 10 g boric acid for preservation. Urine samples were then sub-sampled into smaller aliquots and stored at −80 °C. This study was approved by Chesapeake IRB (Pro00024511) and is registered on clinicaltrials.gov as NCT03783013.

**Figure 2.**
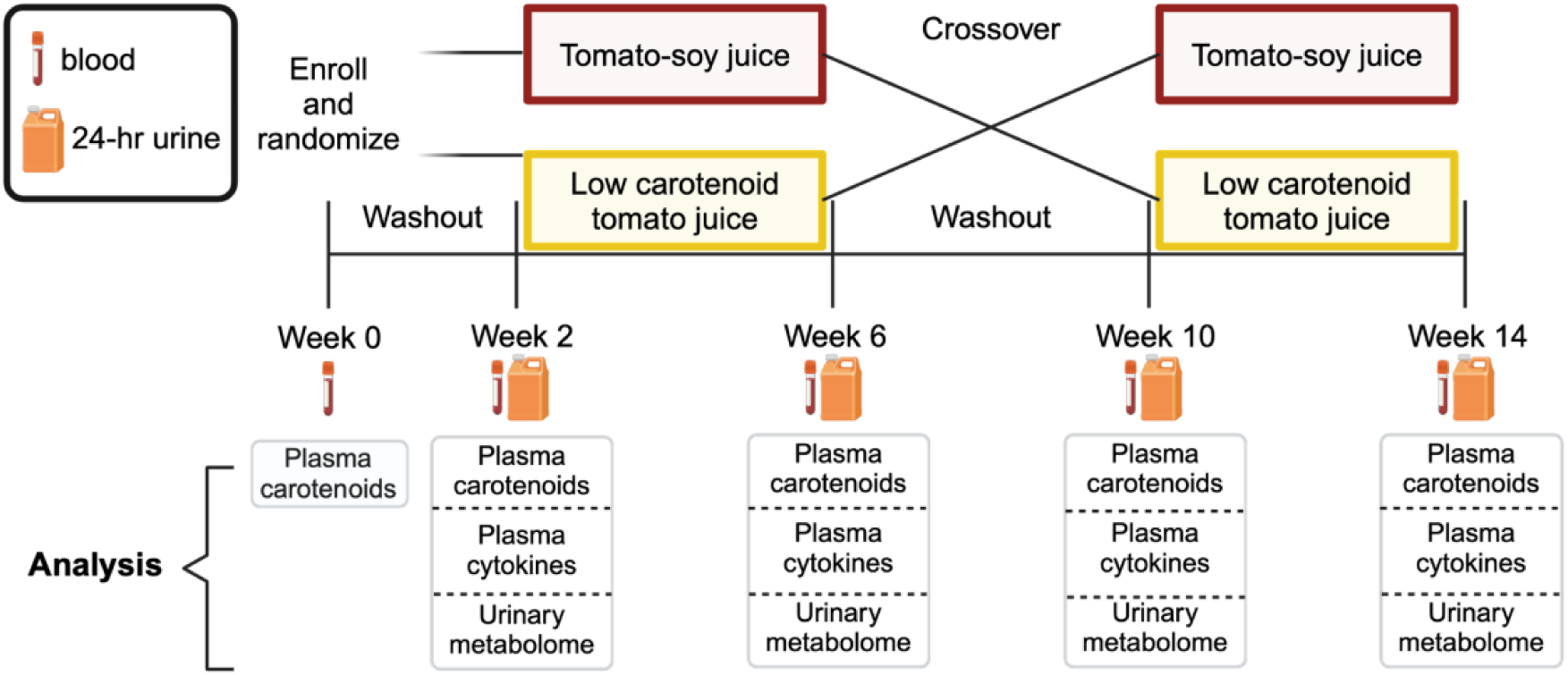
Schematic of crossover clinical trial (clinicaltrials.gov: NCT03783013) providing subjects with obesity a daily intervention of high lycopene tomato-soy juice and control (low carotenoid, no soy) tomato juice.

### 2.2 Study treatments

Two tomato varieties grown at the North Central Agricultural Research Station at Ohio State University (OSU) were used to create juices for this clinical trial. The control tomato juice was made from OH8614, a low-carotenoid experimental inbred hybrid with a non-functional phytoene synthase, an enzyme necessary for carotenoid biosynthesis. The inbred line OH8614 is homozygous for the recessive allele *r* (i.e., yellow flesh, a non-functional phytoene synthase mutation) derived from a cherry tomato variety (PI 114490) and bred into a processing tomato variety (Kang et al., 2014). For the tomato-soy juice, FG99-218, a high-lycopene inbred variety with dark green (*dg*) and old gold crimson (*og^c^*) alleles in *B*, was used. These tomatoes were mechanically harvested, sorted for full ripeness, peeled, and hot break processed into juice at the OSU Food Industries Center, following good manufacturing practices, as we have done previously (Cooperstone et al., 2015). Fully ripened high lycopene tomatoes were processed into juices and additionally enriched with Solgen 40S, a 40% soy isoflavone extract derived from germ (Tradichem, Madrid, Spain), prior to hot break processing, similar to as done previously (Bohn et al., 2013).

### 2.3 Chemicals

β-Carotene, uridine 5ʹ-diphosphoglucuronic acid (UDGPA), magnesium chloride, β-nicotinamide adenine dinucleotide 2′-phosphate reduced tetrasodium salt hydrate (NADPH), Tris hydrochloride, 4-ethylphenol sulfate, 4-ethylphenol, dihydrogenistein, tyrosol, LC-MS Optima grade water, acetonitrile, methanol, ammonium formate, and formic acid were all purchased from Sigma-Aldrich (St. Louis, MO, USA). Ammonium hydroxide and hydrochloric acid were obtained from Fisher Scientific (Waltham, MA, USA). Daidzein, daidzin, genistein, genistin, glycitein, and glycitin were all purchased from ChromaDex (Longmont, CA, USA). O-Desmethylangolensin (O-DMA), 6ʹ-hydroxy-O-DMA, and dihydrodaidzein were obtained from Toronto Research Chemicals (Ontario, Canada). α-Carotene was purchased from Cayman Chemicals (Ann Arbor, MI, USA). β-cryptoxanthin, lutein, and zeaxanthin were purchased from Extrasynthese (Genay, France). 3ʹ-Phosphoadenosine-5-phosphosulfate (PAPS) was obtained from R&D Systems (Minneapolis, MN, USA). Lycopene was extracted and crystallized from tomato paste and spectral purity was determined to be >98% (Kopec et al., 2010).

### 2.4 Determination of phytochemical contents in intervention juices

#### 2.4.1 Soy isoflavone extraction and HPLC-DAD analysis in juices

Both tomato-soy and control tomato juices were analyzed for soy isoflavone contents (Murphy et al., 2002). A 1-g aliquot of each juice was weighed into 12-mL glass tubes containing 4 mL of 60% acetonitrile. Following a cold water-bath sonication for 5 minutes, samples were centrifuged for 10 min at 4,000 × *g*. The aqueous supernatant was transferred to a 25 mL volumetric flask, and the extraction was repeated 3 times more. After bringing up to volume, 1 mL was dried down under nitrogen gas and stored in −80 °C until analysis using high-performance liquid chromatography coupled with diode array detection (HPLC-DAD).

Juice sample extracts were redissolved in methanol followed by bath sonication for 2 minutes. Extracts were filtered (0.22 μm nylon) and transferred to HPLC vials for analysis. On an Agilent 1260 HPLC-DAD, 1 μL of each extract was injected onto a Zorbax Eclipse Plus C18 column (2.1×100 mm, 1.8 μm particle size, Agilent Technologies) using a gradient with water and 0.1% formic acid (A), and acetonitrile and 0.1% formic acid (B). The gradient is as follows: begin at 10% B and hold for 0.5 min, increase linearly to 40% B for 2 min, increase linearly to 45% B for 6.5 min, increase linearly to 55% for 2 min, increase linearly to 100% B over 1 min, and return to initial conditions for 2 min, for a total run time of 14 min. The column was maintained at 30 °C and the flow rate was 1 mL/min. Daidzein, genistein, glycitein, and their glycosylated forms (daidzin, genistin, glycitin) were quantified using an adapted method (Murphy et al., 2002).

#### 2.4.2 Extraction and HPLC-DAD analysis of carotenoids in juices

Carotenoids were extracted from tomato-soy and control juices following a previously described method (Kopec et al., 2010), and analyzed using HPLC-DAD as reported previously (Cooperstone et al., 2015). Using external standard curves, lycopene, β-carotene, α-carotene, zeaxanthin, lutein, and β-cryptoxanthin were quantified. Phytoene and phytofluene were quantified using a ratio of each molar extinction coefficient to lycopene to create a relative slope (Cooperstone et al., 2016).

### 2.5 Assessment of inflammation biomarkers

To determine the effects of the dietary tomato interventions on systemic inflammation, inflammatory cytokines from blood plasma were measured. Fifteen cytokines (interleukin (IL)-6, IL-8, TNF-α, monocyte chemoattractant protein-1 (MCP-1), interferon (IFN)-γ, granulocyte-macrophage colony-stimulating factor (GM-CSF), IL-10, IL-12p40, IL-12p70, IL-13, IL-1β, IL-1rα, IL-2, IL-4, and IL-5) were quantified in duplicate in each sample using a bead based multiplex immunoassay by Eve Technologies (Calgary, AB, Canada).

### 2.6 Extraction and analysis of blood plasma carotenoids

Carotenoid extraction from blood plasma (500 μL) was conducted as previously described (Sholola & Cooperstone, 2022), using the same HPLC-DAD method that was used for juice analyses (Cooperstone et al., 2015).

### 2.7 Urine metabolome profiling

#### 2.7.1 Urine sample preparation

After thawing in cold water, urine was centrifuged for 10 min at 21,130 × *g*. Urine osmolality was measured by freezing point depression using an osmometer (Advanced Osmometer Model 3300; Norwood, MA, USA). To account for differences in urine volumes and hydration level between subjects/timepoints, each sample was normalized to 100 milliosmoles with water. Normalized urine samples were added to new sample tubes containing methanol and acetonitrile, so that the final mixture was the injection solvent 2:1:1 ACN/MeOH/H_2_O and centrifuged at 21,130 × *g* for 5 min to pellet any insoluble material. Process blanks were prepared using water following the same steps. Quality control (QC) samples were prepared by pooling 100 μL from each normalized urine sample.

Samples were analyzed on a 1290 Infinity ultra-HPLC (UHPLC) coupled to a 6546 quadrupole time-of-flight (QTOF) MS with electrospray ionization (ESI) in both positive (+) and negative (-) ionization modes (Agilent Technologies, Santa Clara, CA, USA). For comprehensive metabolite coverage, data were collected for each sample in 4 combinations of analysis: reverse-phase (RP)LC-MS (+), RPLC-MS (-), hydrophilic interaction LC (HILIC)-MS (+), and HILIC-MS (-). For each analysis, sample order was randomized, with QC samples in every 7^th^ position (i.e., approximately every 1.5-2 h) to monitor instrument variability throughout the duration of the experimental run. Three process blanks were queued at the beginning of each run to account for residues and contamination from extraction materials and remove them from downstream analyses.

For RPLC-MS analysis, 20 μL was injected onto a 2.1 x 100 mm x 1.8 μm Waters HSS column (Milford, MA, USA) kept at 40 °C. The mobile phase consisted of (A) 0.1% formic acid in water, and (B) 0.1% formic acid in acetonitrile and the gradient was applied at 0.4 mL/min as described: held at 1% B for 0.5 min, increased linearly to 100% B over 10 minutes, held there for 1 min, then immediately decreased to 1% B and held there for 2 min for a total time of 13.5 min. Full scan data was acquired in a mass range of 50-1700 *m/z* with a scan rate of 3 spectra/sec. MS parameters were set as followed: gas temperature of 350 °C, gas flow at 10 L/min, nebulizer at 35 psig, sheath gas temperature at 375 °C, and sheath gas flow at 12 L/min.

HILIC analysis was adapted from methods in Spagou et al (2011). Twenty μL was injected onto a 2.1 x 100 mm x 1.8 μm Waters Z-HILIC column maintained at 40 °C. Mobile phase consisted of (A) 1:1 acetonitrile/water in 10 mM ammonium formate (pH 7.2) and 0.1% formic acid, and (B) 95:5 acetonitrile/water in 10 mM ammonium formate (pH 7.2) and 0.1% formic acid. The 14 min gradient was applied at 0.5 mL/min as described: held at 100% B for 1 min, increased linearly to 100% A over 10 min and held at this point for 1 min. Immediately afterwards, the gradient was held at 100% B for 2 min. MS source parameters were set to the same parameters described for RPLC-MS analysis.

### 2.8 Data analysis

#### 2.8.1 Univariate statistical analyses of carotenoids and inflammatory cytokines

Baseline characteristics for participants were summarized using means and standard deviations. Changes in carotenoid levels and inflammatory proteins in blood plasma samples were initially investigated using linear mixed effect models to incorporate the crossover design and intervention order. Several models included variables such as sex and age, and were tested to adjust for potential contributions to outcomes. The best mixed effect model was selected based on the lowest corrected Akaike’s information criterion. Ultimately, carotenoids (Student’s *t* tests) and cytokines (Wilcoxon rank-sum tests) with *p* < 0.05 were considered statistically significant. For missing values in the cytokine data (detected below the limit of quantification), the values were imputed with half of the lowest concentration for each protein.

#### 2.8.2 Metabolomics data preprocessing and statistical analyses

Spectral data was converted into a list of chemical features and their abundances using MZmine3 (Schmid et al., 2023). Raw spectral data files obtained from UHPLC-QTOF-MS analysis were converted from Agilent .d files to .mzML files using ProteoWizard’s MSConvert (Chambers et al., 2012) and imported into MZmine3 for data deconvolution. Feature extraction and alignment parameters can be found in **Table S1, Supplementary Materials**. Data filtering and analysis was conducted in R v4.4.1. To ensure data quality, features in each metabolomics dataset were filtered based on the following criteria: must be present in every QC, must have a coefficient of variation less than 30% across the QCs, and intensities must be at least 10x higher in QC than in process blanks. Missing values were imputed by replacing them with half of the lowest peak intensity of each feature. Each dataset was log_2_-transformed after filtering and imputation. Intensity drifts were then corrected using the *notame* package in R (Klåvus et al., 2020) (**Figures S1-S2, Supplementary Material**), and similar features (at least 90% correlation and within 1 second retention time window) were clustered to reduce redundancy using the same package. Principal component analyses were used for data visualization with *FactoMineR* (Lê et al., 2008) and *mixOmics* (Le Cao et al., 2018) packages. Univariate statistics such as paired *t*-tests and analysis of variance (ANOVA) were applied to all metabolomics datasets and corrected for multiple testing using the Benjamini-Hochberg procedure with the *rstatix* package.

#### 2.8.3 Metabolomic feature identification

Data-dependent and independent UHPLC-QTOF-MS/MS experiments were conducted on pooled treatment samples in all 4 combinations of analysis to aide in identification of features. MS2 experiments were conducted using similar parameters as MS1 except collision energies of 20 eV and 40eV, an MS1 scan rate of 4 spectra/sec, and an MS2 scan rate of 2 spectra/sec were used. Authentic or bio-synthesized (Cuparencu et al., 2016) standards were used to further identify some features by comparing accurate mass, retention time, and fragmentation patterns collected from standards to peaks of interest. Annotated masses are detailed in **Tables S2-S3, Supplementary Material**. Confidence levels for identified compounds were adapted from guidelines proposed by the Metabolomics Standards Initiative (Sumner et al., 2007). In short: level 1 is a compound identified based on both retention time and *m/z* match to authentic standard; level 1.5 is a compound identified based on retention time and *m/z* match to bio-synthesized standard; level 2 identified compound is based on mass spectra matches to spectra published in the literature; level 3 is when compound class is identified; and level 4 is unknown (i.e., just *m*/*z* and retention time reported).

#### 2.8.4 In vitro biosynthesis

##### 2.8.4.1 Metabolite biosynthesis from standards

To confirm the identification of glucuronidated, sulfonated, and oxidized metabolites, S9 pig liver fractions and cofactors were incubated with precursor standards (i.e., bio-synthesized) following methods adapted from Cuparencu et al (2016). Hepatic S9 isolates were prepared from pig liver as described in Wu and McKown (2004). Metabolites were synthesized by incubating 10 μL standard solution, 10 μL of S9 liver enzymes, and the following cofactors in Tris hydrochloride buffer: 10 μL of 20 mM UDGPA, 12.5 μL of 40 mM NADPH, 10 μL of 2 mg/mL PAPS, and 10 μL 100 mM magnesium chloride. Samples were dried down using an Eppendorf vacufuge (Hamburg, Germany) and reconstituted in the same injection solvent used for urine metabolomics analysis (2:1:1, ACN/MeOH/H_2_O). To confirm the presence of synthesized metabolites from the in vitro enzyme system, bio-synthesized sample, fresh standard, and two controls: S9 liver enzymes and S9 liver enzymes with cofactors, were injected onto the UHPLC-QTOF-MS system following the method described in **Section 2.7.2**.

##### 2.8.4.2 Metabolite biosynthesis from intervention juices

To further validate diet-derived metabolite annotations, the same biosynthesis approach and LC-MS analysis described in **Section 2.8.4.1**. was applied to hydrolyzed extracts of intervention juices. These samples were injected and analyzed alongside bio-synthesized metabolites, fresh standards, and controls.

Acid hydrolysis was performed on juice extracts (from both control and tomato-soy) to mimic enzymatic hydrolysis of glycosylated phytochemicals in vivo. Using a method adapted from Chiang et al. (2001), 3 N HCl in methanol was added to 1.5g tomato-soy or control juice, sonicated for 15 mins, and incubated in a shaking water bath for 4 hours at 70 °C and 70 RPM. After the hydrolyzed extract was neutralized with 3 N ammonium hydroxide and centrifuged for 5 mins at 4,000 × *g*, supernatants were filtered (0.22 μm nylon) and analyzed for masses of interest from extracted ion chromatograms.

### 2.9 Dataset availability

Raw MS1 data are available in Metabolomics Workbench (Sud et al., 2016) (ST004178) and data-dependent MS/MS data have been reposited with GNPS’s MassIVE (<u>MSV000099128</u>) (M. Wang et al., 2016). Code for metabolomics, carotenoids, and cytokines analyses can be found at https://github.com/CooperstoneLab/TomatoSoy-Obesity-Metabolomics.

## 3 Results and Discussion

### 3.1 Characterizing intervention agents allow for plausible links between bioactive compounds and health effects while enabling cross-study comparisons

Tomato-soy juice provided in this study was intended to deliver doses of tomato lycopene and soy isoflavones that have been associated with reduced cancer risks in observational studies (Giovannucci et al., 1995, 2002; Shu et al., 2009). A team of Ohio State researchers previously demonstrated that carotenoids and soy isoflavones are readily absorbed from tomato-soy juice in a phase I clinical study, achieving levels comparable to populations at lower risks for chronic diseases (Bohn et al., 2013). A few years later in another tomato-soy intervention, the same team showed that lycopene and soy isoflavones elicit dose-response increases in the plasma and urine, respectively, in prostate cancer patients who consume 1 can (21 mg lycopene, 66 mg soy isoflavones) or 2 cans of tomato-soy juice daily (Grainger et al., 2019). In this study, the tomato-soy juice provided 54 mg/lycopene and 189.9 mg/total isoflavones per day. The quantities of individual carotenoids and soy isoflavones provided in the intervention juices for this study are listed in **Table 2**, where lycopene is the predominant carotenoid in tomato-soy, and genistin is the most abundant soy isoflavone.

**Table 2.** Average carotenoid and soy isoflavone contents in tomato-soy and low carotenoid tomato juice interventions (± standard deviation). Results are the average of n = 5 replicates per juice.

|  | Tomato-Soy<br>Juice | Control, Low Carotenoid<br>Tomato Juice |
| --- | --- | --- |
| **Carotenoid, mg/d** |  |  |
| Beta-carotene | 1.8 ± 0.33 | 0.2 ± 0.02 |
| Lycopene | 54 ± 9.30 | 1.5 ± 0.12 |
| Phytoene | 12.6 ± 2.13 | < LOQ |
| Phytofluene | 4.6 ± 0.77 | < LOQ |
| Total carotenoids | 73.0 ± 12.5 | 1.7 ± 0.14 |
| **Soy isoflavone, mg/d** |  |  |
| Daidzin | 26.9 ± 2.3 | n.d. |
| Glycitin | 17.1 ± 2.3 | n.d. |
| Genistin | 142.6 ± 20.0 | n.d. |
| Daidzein | 0.9 ± 0.2 | n.d. |
| Glycitein | 0.5 ± 0.1 | n.d. |
| Genistein | 1.9 ± 0.1 | n.d. |
| Total soy isoflavones | 189.9 ± 21.6 | - |

< LOQ, below limit of quantification

Daily lycopene intake of up to 21 mg has been inversely associated with prostate cancer risk (P. Chen et al., 2015). In a typical American diet, lycopene intake is variable, ranging from about 3-15 mg/d based on epidemiological evidence (Porrini & Riso, 2005). A standard serving of tomato sauce contains approximately 10-17 mg per serving (Sholola & Cooperstone, 2022), whereas participants in this study were administered ∼54 mg of lycopene from the tomato-soy juice intervention–exceeding what is typically consumed in Western diets and comparable to what has been previously provided to prostate cancer subjects consuming 2 cans of tomato-soy juice/day (42 mg lycopene) (Grainger et al., 2019). Phytoene and phytofluene were quantifiable only in the tomato-soy juice, averaging to 12.6 and 4.6 mg per day, respectively. Phytoene and phytofluene levels were too low to be quantified in the control tomato juice. The control juice was made using yellow-flesh tomatoes that contain a non-functional phytoene synthase enzyme (see **Materials and Methods, Section 2.2**.). This enzyme is the rate-limiting step of carotenoid biosynthesis and dimerizes two geranylgeranyl pyrophosphate molecules to form phytoene, which is converted to phytofluene, and subsequently other carotenoids are produced downstream (Gady et al., 2012). Due to the phytoene synthase mutation in the yellow tomatoes used for the control, this juice was expected to reflect very low levels of carotenoids, as observed (**Table 2**). These carotenoids though are not completely absent, as carotenoid production in tomatoes is controlled independently in the fruit flesh and peel, and *r* is only present in the flesh (BUTLER, 1952; Fray & Grierson, 1993, 1993).

Cancer incidence, particularly breast cancer, is lower in East Asian countries (e.g., Japan and China), a difference that is often attributed to soy-rich diets (He & Chen, 2013). While adults in Japan consume up to 30-50 mg soy isoflavones/day, intake in Western countries (where less soy is eaten) is substantially lower at less than 3 mg/day (Messina, 2016). In this study, participants received an average of 189.9 mg total soy isoflavones/day, of which ∼98% were glycosides (daidzin, glycitin, and genistin). Genistin made up a majority of isoflavone content at an average of 142.6 mg/day (75% total isoflavones). Grainger et al. (2019) also provided similar amounts of total isoflavones (130 mg) to prostate cancer subjects consuming 2 cans of tomato-soy juice daily, with genistein equivalents making up 65% of the total isoflavones. Aglycones (daidzein, glycitein, and genistein) made up only a small amount (∼0.02%) of total isoflavone content here, which is expected for unfermented soy-containing foods, as at least 90% isoflavones exist as glycosides (H. Wang & Murphy, 1994b, 1994a).

As mentioned in the beginning of this section, this tomato-soy intervention was designed to maximize the contents of lycopene and soy isoflavone components co-administered in a whole food product. By characterizing the intervention juices, we lay a foundation for understanding the metabolic and physiological impacts of high lycopene and soy isoflavone intake while facilitating meaningful comparisons across literature.

### 3.2 Carotenoids significantly increase in blood plasma only after subjects consume tomato-soy juice

To investigate changes in blood plasma carotenoid levels, linear mixed effect models were first applied. There were no sequence carry-over effects caused by the previous intervention, and sex and age did not contribute to carotenoid levels (*P* > 0.05). Therefore, paired Student’s *t* tests were used to determine significant changes. Total lycopene and β-carotene levels were significantly increased overall in blood following tomato-soy consumption, while the same subjects mostly showed unchanged or decreased levels during the control, low-carotenoid tomato intervention (Figure 3). Only one subject exhibited increased total lycopene levels (Figure 3) following the control intervention, suggesting noncompliance and was thus excluded from downstream analyses when making pre- to post-control and post-control to post-tomato-soy comparisons. At baseline (week 0), we observed subjects to have ∼450 ± 293 and 950 ± 380 nmol/L of plasma β-carotene and lycopene, respectively (**Table S4, Supplementary Material**). This is in line with typical plasma carotenoid levels that are found in Americans, averaging ∼300 nmol/L β-carotene and ∼800 nmol/L lycopene (Centers for Disease Control and Prevention (CDC), 2005; Sholola & Cooperstone, 2022).

**Figure 3.**
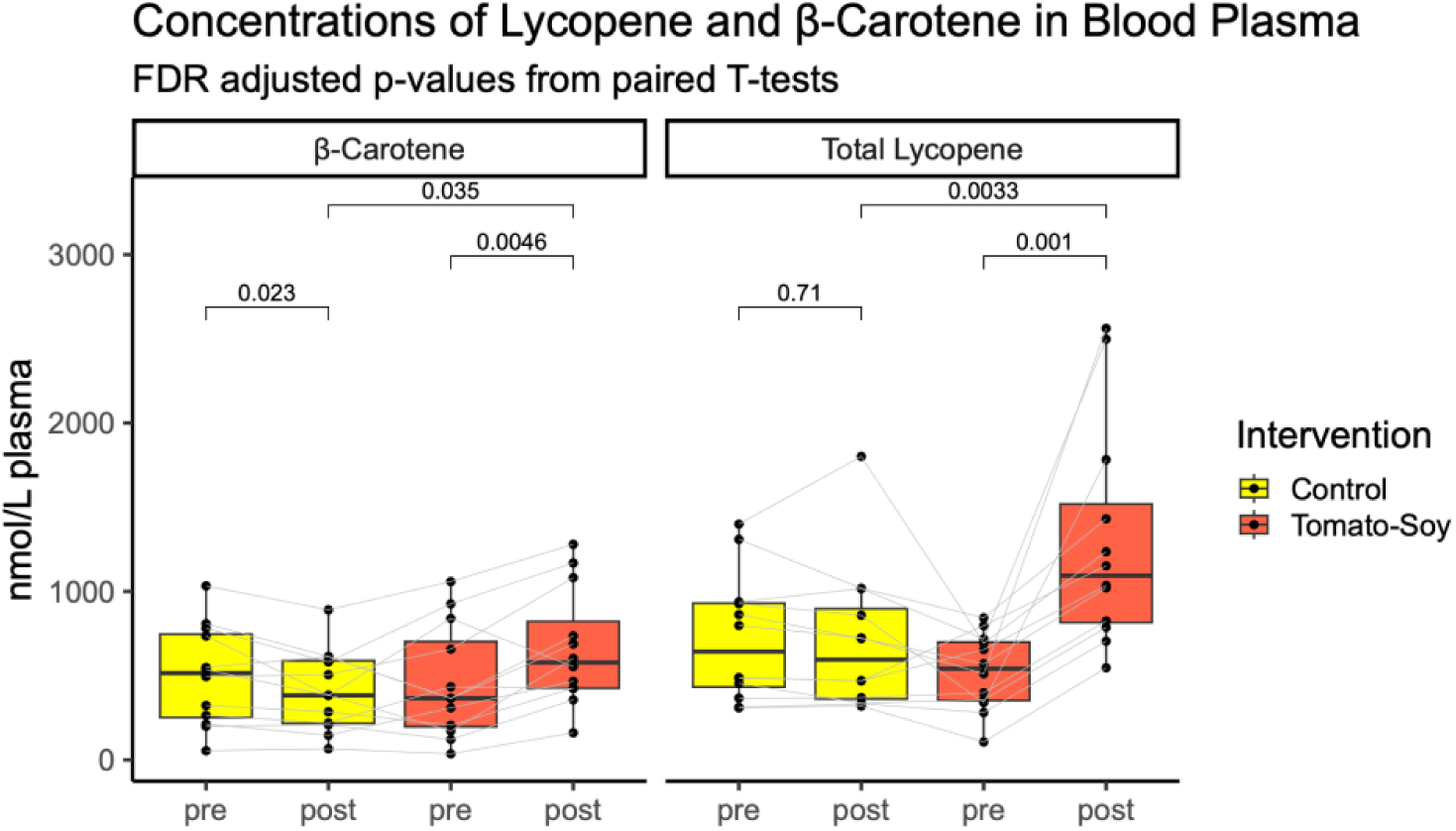
Carotenoid levels in blood plasma before and after control and tomato-soy interventions. Each point represents one subject, and lines connect the points for the same subject.

Given that lycopene was the predominant carotenoid in tomato-soy juice, it was expected to be the most abundant carotenoid measured in blood plasma post-intervention. We found that lycopene levels reached 1,298.4 (± 664.71) nmol/L after tomato-soy juice intake, an average increase of 2.82-fold from pre-intervention. All subjects exhibited an increase of circulatory lycopene with tomato-soy consumption. Lycopene levels did not significantly change when subjects consumed the low-carotenoid control, though they did numerically decrease by about 7%, excluding the outlier subject (Figure 3). Plasma lycopene was significantly higher (2.18-fold) at the end of the tomato-soy intervention as compared to post-control juice (Figure 3).

Similar trends were observed for β-carotene following tomato-soy intake. Plasma β-carotene levels increased significantly to 662.1 (± 347.95) nmol/L, a 2-fold average increase when compared to pre-tomato-soy (Figure 3, **Table S4, Supplementary Material**). Following control juice consumption, β-carotene levels significantly decreased by ∼15% when compared to before intake. By the end of the tomato-soy intervention, plasma β-carotene levels were significantly higher (1.86-fold) when compared to post-control juice consumption (Figure 3). Additionally, phytofluene levels were significantly increased from ∼200 to 350 nmol/L following tomato-soy while being significantly decreased from 975 nmol/L to ∼240 nmol/L after control (**Table S4, Supplementary Material**). The remaining carotenoids quantified (phytoene, α-carotene, β-cryptoxanthin, lutein, and zeaxanthin) in the blood remained unchanged in both interventions (**Table S4, Supplementary Material**).

The levels we report post-tomato-soy intervention are consistent with what has been previously reported for tomato-based interventions, where plasma lycopene concentrations often exceed 1,000 nmol/L following daily intake. For example, in type 2 diabetes patients, plasma lycopene levels increased to a mean concentration of 1,008 nmol/L while CRP levels decreased by the end of a 4 week intervention drinking 500 mL tomato juice a day (Upritchard et al., 2000). In the first pilot study utilizing tomato-soy juice (22 mg lycopene/day), it was found that plasma lycopene concentrations can accumulate to 1,280 nmol/L on average after 4 weeks of consumption in healthy men and women (Bohn et al., 2013). By the end of the study (week 8), lycopene concentrations in plasma were found to be similar (∼1,240 nmol/L), and lipid antioxidant status was significantly improved (Bohn et al., 2013). It has also been shown that after 3 weeks of tomato-soy juice consumption (2-can dose, ∼40 mg lycopene) in men with prostate cancer, plasma lycopene levels accumulated to ∼1900 nmol/L on average (Grainger et al., 2019), and was significantly correlated with prostate lycopene concentrations (average of 0.67 nmol/g following 2-can dose) (Grainger et al., 2019). Daily tomato sauce consumption providing 30 mg lycopene resulted in plasma levels of ∼1300 nmol/L in men with prostate tumors, and was associated with increased apoptosis in both benign and malignant tumor biopsies (Kim H et al., 2003). In another human study, subjects consumed tomato-based products providing 30 mg lycopene every day for 4 weeks, increasing average serum lycopene to ∼685 nmol/L, while also significantly reducing oxidative stress (Rao, 2004). Additionally, in vitro studies demonstrate that lycopene concentrations between 500-2,000 nmol/L can inhibit oxysterol-induced pro-inflammatory responses in human macrophages (Palozza et al., 2011). Previous studies suggest that circulating levels achieved in the present study are sufficient to elicit health-protective effects (Bohn et al., 2013; Kim H et al., 2003; Palozza et al., 2011; Rao, 2004).

As evidenced by large standard deviations for carotenoids measured here (**Table S4, Supplementary Material**), many factors can impact carotenoid metabolism, distribution, absorption, and thus response levels in the blood. Dietary factors such as the food matrix and co-consumed components can significantly impact absorption. Lycopene is more efficiently absorbed from heat-processed tomato products when compared to raw tomatoes (Gärtner et al., 1997; Porrini et al., 1998), and as noted in **Section 3.1**, the bioavailability of carotenoids from tomato-soy juice has been previously characterized (Bohn et al., 2013). While the use of a processed product in this study provided a standardized matrix for consumption and facilitated optimal lycopene bioavailability, participants were not instructed to consume tomato-soy juice with specific foods. This may partly explain the variability in carotenoid levels observed in this work, as co-ingestion of carotenoids with fats can enhance absorption (Brown et al., 2004; Kopec et al., 2014; Unlu et al., 2005), whereas fiber could inhibit it (Horvitz et al., 2004; Riedl et al., 1999; Rock & Swendseid, 1992). Notably, numerous candidate gene and genome-wide association studies show that genetic variation can also influence circulating carotenoids in people (Beydoun et al., 2014; Borel et al., 2011; D’Adamo et al., 2016; Farook et al., 2017; Ferrucci et al., 2009; Moran et al., 2018; Zubair et al., 2015), with variants in genes such as *SCARB1* (scavenger receptor class B member 1) and *BCO1* (β-carotene oxygenase 1) frequently associated with plasma lycopene and β-carotene concentrations (Borel, 2012; Borel et al., 2015; Ferrucci et al., 2009; Hendrickson et al., 2012; Moran et al., 2019; Zubair et al., 2015). A previous study in prostate cancer patients consuming the same tomato-soy intervention found that single nucleotide polymorphisms (SNPs) in *BCO1* were significant predictors of changes in plasma lycopene and β-carotene levels (Moran et al., 2019), further emphasizing the role of genetic variation for circulating carotenoids. These associations are increasingly documented, with carotenoid status also being influenced by extrinsic factors such as body weight, lifestyle, and age that have been thoroughly reviewed (Bohn et al., 2017; Moran et al., 2018). The crossover design of this study was intended to account for and capture interindividual variation in carotenoid responses.

### 3.3 Pro-inflammatory cytokines are reduced in blood plasma only while subjects are on tomato-soy intervention

To measure the modulatory effects of lycopene and soy isoflavones administered from tomato-soy juice on low-grade systemic inflammation, soluble markers in blood plasma were assessed before and after each intervention. Sequence effects (i.e., the other of the intervention), sex and age did not contribute to cytokine response levels (*P* > 0.05) in mixed linear models; thus, Wilcoxon rank-sum tests were used to investigate differences since Q-Q plots demonstrated non-normal data. Plasma cytokine levels for each subject (along with carotenoid levels) can be found in **Table S5, Supplementary Material**. A significant reduction of 3 pro-inflammatory cytokines (GM-CSF, IL-12p70, and IL-5) were observed when comparing levels pre- and post-tomato-soy intake (Figure 4). TNF-α also was trending downwards, and nearly reached significance (P = 0.052). These reductions were not observed when comparing post-tomato-soy to post-control, or pre-control to post-control. This pattern suggests that tomato-soy intake elicits an anti-inflammatory response. The absence of the same observations in post-control vs. post-tomato-soy comparisons indicates some complexity in determining whether these effects are driven by lycopene, soy isoflavones, and/or other phytochemicals present in the tomato-soy juice matrix. Several studies have demonstrated the anti-inflammatory effects (e.g., reduction of pro-inflammatory cytokines: TNF-α, MCP-1, IFN-γ, IL-6, and IL-8) of tomato alone (Ghavipour et al., 2013; Y.-F. Li et al., 2015; Tsitsimpikou et al., 2014) and soy alone (Ahn-Jarvis et al., 2020; Azadbakht et al., 2007; Lesinski et al., 2015; Llaneza et al., 2011) in subjects with elevated inflammatory statuses (i.e., obesity/overweight, metabolic syndrome, prostate cancer, chronic pancreatitis). We selected to use a low-carotenoid tomato as our control to isolate the effect of lycopene and isoflavones, separate from other tomato-derived bioactives. Based on these studies, the sample size of 30 participants for this clinical trial was determined a priori (α=0.05, 80% power) using G*Power 3.1 (Faul et al., 2007) to primarily detect changes in TNF-α, MCP-1, IFN-γ, IL-6, and IL-8 levels. However, due to the disruption of the COVID-19 pandemic, it was no longer feasible to reach the desired enrollment of at least 30 participants at the time. A significant response was not observed for the hypothesized pro-inflammatory cytokines as expected, but due to a low sample size we cannot determine that there is no effect after tomato-soy intake. Using the same alpha and beta, and conducting a post-hoc power calculation for IL-6 concentrations (using Wilcoxon ranked test given the non-normal data), we would have needed 108 individuals to see a difference between post-tomato-soy vs. post-control tomato juice. The same rationale can be applied to the lack of statistically significant observations when comparing post-control to post-tomato-soy; particularly for pro-inflammatory cytokines significantly reduced following tomato-soy juice intake (i.e., GM-CSF, IL-12p70, IL-5).

**Figure 4.**
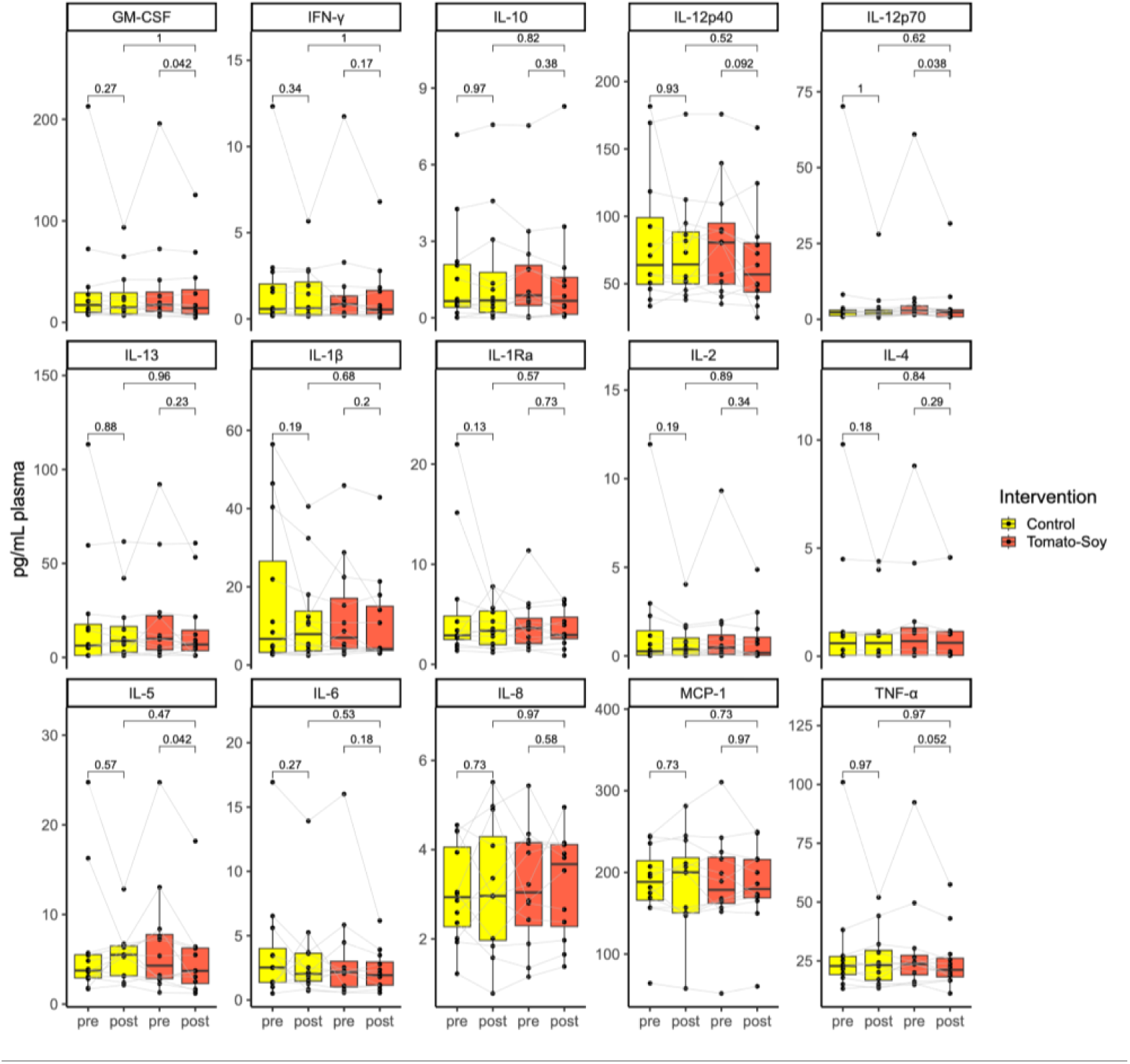
Pro-inflammatory cytokines measured before and after control and tomato-soy intervention (n = 11 or 12) interventions using bioplex-based assay. False discovery rate corrected P-values from Wilcoxon signed-rank tests. Significance is defined by *P* < 0.05

Like other T helper 2 (Th2)-cell associated cytokines, IL-5 is closely linked to asthmatic inflammation (P. J. Barnes, 2018; Hogan et al., 1997; Hogan & Foster, 1996; Lambrecht et al., 2019). IL-5 is reported to be elevated in subjects with general or abdominal obesity when compared to individuals categorized as normal-weight, while also positively correlating with fractional exhaled nitric oxide (a biomarker for airway inflammation) (Salem, 2022). Consistent with this study, work from our groups have shown in a mouse model of chronic pancreatitis that tomato-soy intake significantly reduces systemic IL-5 (Mukherjee et al., 2020). Mukherjee et al. (2020) hypothesized that the reduction in IL-5 seen in mice with chronic pancreatitis could be an indicator that tomato-soy intake could alter eosinophil production, since eosinophils can drive IL-5 secretion (Gitto et al., 2020; Manohar et al., 2018) and eosinophilia has been observed in patients with chronic pancreatitis (Q. Wang et al., 2009). Interestingly, soy isoflavones (Bao et al., 2011) and lycopene (Hazlewood et al., 2011) have separately been shown to attenuate IL-5 levels and airway inflammation in murine models of allergic asthma. In a cross-species study, Quail et al. (2017) demonstrate that elevated serum GM-CSF expression in both high fat diet-induced obese mice contributes to inflammatory lung conditions by promoting lung neutrophilia. The same study included a secondary analysis showing that a 10% weight loss through caloric restriction in obese females led to systemic reductions of IL-5 and GM-CSF (Alemán et al., 2017; Quail et al., 2017). In mice with diet-induced obesity, IL-12p70 levels were significantly higher than in mice fed low fat diets (Hildreth et al., 2023). IL-12 induces the secretion of several cytokines (e.g., GM-CSF and TNF-α) from T-cells and natural killer (NK) cells (Chan et al., 1991; Kobayashi et al., 1989; Kubin et al., 1994; Trinchieri, 2003). In obese adipose tissue, TNF-α is overexpressed, contributing to systemic inflammation in obesity (Hotamisligil et al., 1993). In our study, TNF-α plasma levels were reduced, though not statistically significant, and may further reflect the dampening effects of tomato-soy on pro-inflammatory signaling. Tomato-soy diets fed in the chronic pancreatitis mouse model mentioned earlier significantly reduced circulatory TNF-α levels (Mukherjee et al., 2020).

Our data show that 4 weeks of tomato-soy juice consumption reduces IL-12p70, IL-5, GM-CSF levels, with the reduction in TNF-α trending towards statistical significance (*P* = 0.052). Collectively, these findings suggest that consuming lycopene/tomato juice and soy isoflavones could play a role in the reduction of pro-inflammatory cytokines in obesity. Each pro-inflammatory cytokine reduced in this study is transcriptionally regulated, at least in part, by the NF-κB signaling pathway (Levine & Levine, 2006; J. Liu & Beller, 2003; Schreck & and Baeuerle, 1990; Takatsu, 2011). Taken together, these observed reductions could suggest that tomato-soy juice alters inflammation through attenuation of the NF-κB pathway, a mechanism previously reported for lycopene and isoflavone supplementation in obesity models (Fenni et al., 2017; Gouranton et al., 2011; Shen et al., 2019). However, inflammatory mechanisms are highly interconnected, involving cytokines, immune cells, and signaling pathways that influence one another in complex ways. This overlapped with the multifactorial nature of obesity limits the ability to draw definitive conclusions from human studies like this. Further research is needed to unravel these complexities.

### 3.4 Tomato juice interventions alter the urinary metabolome, with tomato-soy driving distinct metabolic changes primarily via soy isoflavone metabolites

#### 3.4.1. Untargeted metabolomics data was suitable for analysis

Metabolomics features after pre-processing and filtering from data collected in RPLC-MS (+/-) and HILIC-MS (+/-), were retained for downstream data analysis. Instrument reproducibility and high data quality is demonstrated by close clustering of QCs in principal components analysis (PCA) plots and log_2_-transformed data quality boxplots (**Figures S3-S4, Supplementary Material**). After feature clustering using the *notame* package in R, metabolomics datasets yielded a total 628 features in RPLC-MS (+), 1409 in RPLC-MS (-), 924 in HILIC-MS (+), and 1,826 in HILIC-MS (-) (see **Table S6-8, Supplementary Material**).

#### 3.4.1 Key urinary features were selected based on response to tomato juice interventions

To find metabolites that were affected by isoflavone or lycopene administration, features needed to be significantly different (*P* < 0.05 using a false discovery rate (FDR)-multiple testing correction) and exhibiting a fold change ≥ 1.5 between pre- and post-tomato-soy, in addition to being significantly different (FDR-corrected *P* < 0.05) between the post-tomato-soy and post-control timepoints (fold change ≥ 1.5). Features that changed in the same direction for both tomato-soy and control interventions were excluded, while those that changed in opposite directions (e.g., increased in control but decreased in tomato-soy) were retained. This resulted in a list of 53 features from RPLC-MS (-), 22 in RPLC-MS (+), 89 in HILIC-MS (-), and 16 in HILIC-MS (+) (see **Table S2, Supplementary Material**).

To discover metabolites in urine influenced by the consumption of tomato juice overall (independent of lycopene and soy), features were selected if significantly increased/decreased (FDR-corrected *P* < 0.05, fold change ≥ 1.5) in the same direction following both tomato interventions. In other words, features needed to be significantly changed in the same direction in both pre-vs. post-tomato-soy, and pre-vs. post-control tomato juice comparisons. Filtering down to key features altered by tomato intake resulted in a list of 3 features in RPLC-MS (-), 3 in RPLC-MS (+), 23 in HILIC-MS (-), and 12 in HILIC-MS (+) (see **Table S3, Supplementary Material**). Identification efforts were prioritized for these selected key urinary features in all four metabolomics datasets.

#### 3.4.2 Bio-synthesized standards are a useful tool for confirming the identities of diet-derived urinary metabolites

Urine metabolomics provides a sensitive assessment for acute responses to dietary intake, often capturing more nutritional-derived components than blood plasma (Walsh et al., 2006, 2007). Phytochemicals such as isoflavones and other phenolics undergo biotransformation by both the host and microbes before excretion as water-soluble derivatives in the urine. While the detection of diet-derived compounds is enhanced in the urine, physiological modifications make their identification challenging, as library matches and standards are more readily available for parent compounds rather than their metabolic products. The use of “bio-synthesized” standards addresses this challenge by providing additional chemically verified reference compounds derived from diet, enabling more confident metabolite annotations.

While plants synthesize flavonoids and conjugate them to sugars to make them less reactive (Cuyckens & Claeys, 2004; Harborne & Mabry, 1982), most flavonoids (including isoflavones) must undergo sugar hydrolysis in the small intestine (by bacterial or mucosal enzymes) to allow absorption in mammals (Day et al., 1998; K. D. R. Setchell et al., 2002). Aglycones then undergo phase II metabolism, are excreted in bile, deconjugated/metabolized by intestinal microflora, and subsequently reabsorbed, entering enterohepatic cycling before excretion as glucuronide or sulfonated conjugates (S. Barnes, 2010; Chang & Nair, 1995). To probe our untargeted data for as many isoflavone metabolites as possible, a list of hypothesized masses derived from isoflavone glycosides (i.e., genistin, daidzin, glycitin) provided in tomato-soy juice following deglycosylation and intestinal/liver metabolism was compiled. O-DMA, for example, is a microbial metabolite of daidzein (Gaya et al., 2016) that has been found as glucuronide and sulfonated conjugates in people (Adlercreutz et al., 1993). An authentic standard of O-DMA was subjected to phase II metabolism in vitro. From the biosynthesized sample, we were able to confirm the identities of two O-DMA glucuronide isomers detected in urine (Figure 5). Extracted ion chromatogram (EIC) peaks which align at a retention time of ∼4.8 min for biosynthesized standard and pooled urine sample are denoted with an asterisk, and the corresponding fragmentation spectra further illustrates a match (Figure 5). Fragment *m/z* 257 appears as the base peak for both spectra, consistent with the neutral loss of glucuronic acid (Levsen et al., 2007) and resulting in the parent compound O-DMA (theoretical *m/z* 257.0819 [M-H]). Fragments *m/z* 175 and 113 (present in both spectra) also correspond to negative mode fragments of glucuronide conjugates that have been previously reported (Holčapek et al., 2008). Additionally, in the literature, *m*/*z* 257 and 175 are reported for O-DMA glucuronide isomers detected in rat urine (Fang et al., 2002). The peaks associated with *m/z* 433 did not appear in EICs for in vitro control samples, as seen in Figure 5, indicating a successful negative control. Metabolites identified in this way we are considering to be “level 1.5” as they do not quite meet the requirement of a match to authentic standards (level 1), but are more conclusively matched with retention time beyond a MS/MS fragmentation pattern (level 2) (Sumner et al., 2007).

**Figure 5.**
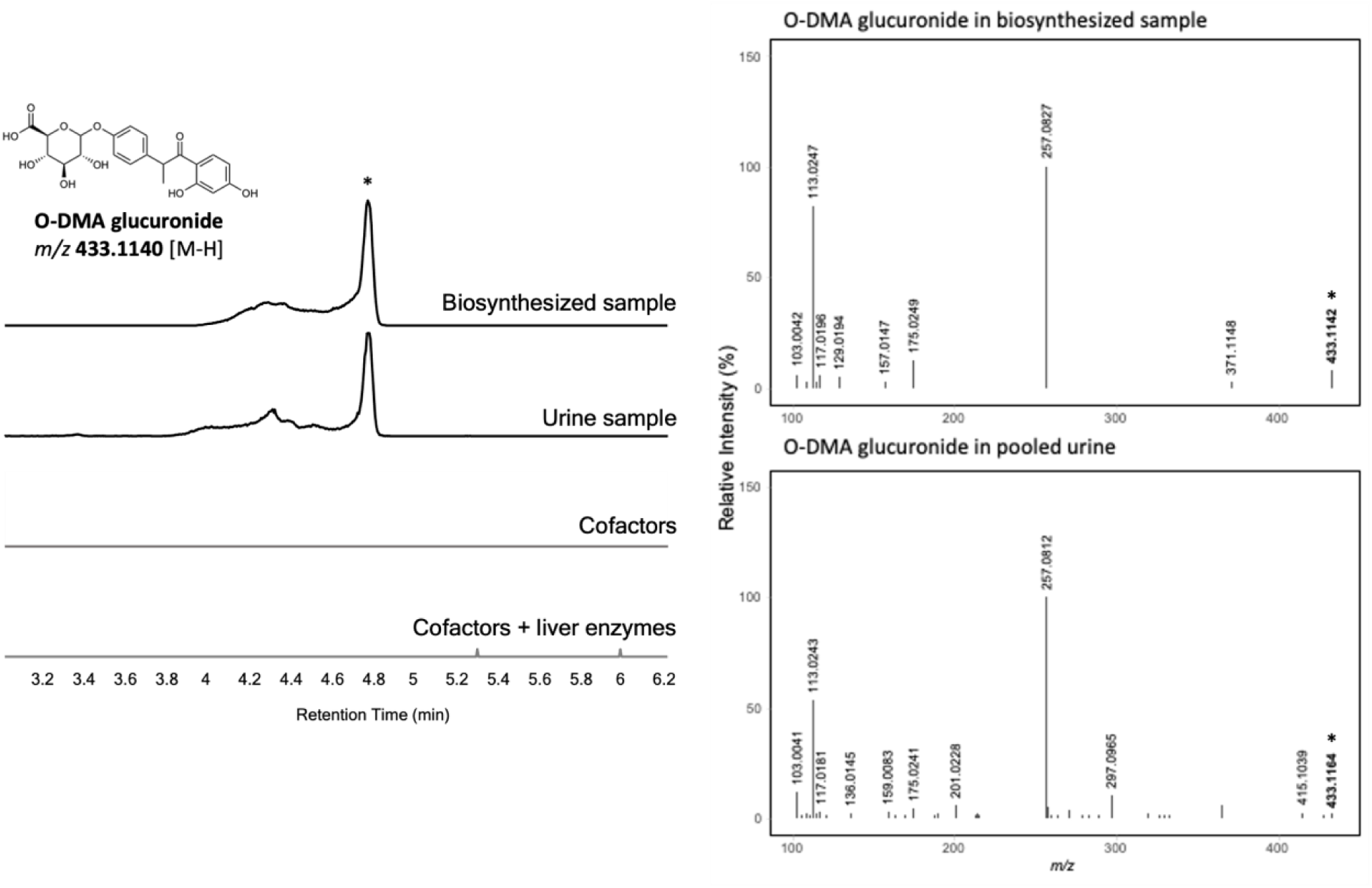
Extracted ion chromatograms (EICs) of *m/z* 433.1140 (O-DMA glucuronide, tentative structure pictured) from biosynthesized standard, urine sample, and control samples (left); MS/MS spectra corresponding to peaks labeled with asterisk in EICs for biosynthesized and pooled urine samples (right)

Biosynthesis methods were also applied to hydrolyzed tomato juices to provide additional reassurance for metabolite annotations. By comparing chromatographic and spectral data from authentic standards of parent compounds, bio-synthesized products, and urine samples, we validated the presence of many key diet-derived features (many of them soy isoflavone metabolites) detected in this study (refer to features labeled with level 1.5 annotation in **Tables S2-S3, Supplementary Material**). With this technique, we annotated 50 metabolites (41 in RPLC-MS and 9 in HILIC-MS datasets).

#### 3.4.3 Tomato-soy intervention alters the urine metabolome, primarily driven by soy isoflavones and their metabolites detected via RPLC-MS (-)

Unsupervised and univariate tests were carried out to determine metabolomic differences in the urine after each intervention timepoint. Among the metabolomics datasets, RPLC-MS (-) revealed the most pronounced group differences (as determined by separation in PCA and number of significantly different features), where urine reflected a distinct metabolic change in urine for post-tomato-soy. This is illustrated in Figure 6A, where PCA demonstrates evident clustering of urine samples at the post-tomato-soy timepoint (dark-red) along PC2, separate from all other treatments/times. The top 5 metabolites driving this separation were ethylphenol conjugates and an O-DMA glucuronide isomer (Figure 6A), products of intestinal metabolism from genistein and daidzein, respectively. Features that were significantly different via repeated measures one-way ANOVA analysis across timepoints are visualized in the heatmap in Figure 6B, with both features and samples organized by hierarchical clustering. This figure is consistent with soy isoflavone metabolites being the primary differentiators in this dataset as they account for 26/54 of these significant differences across groups. Urine samples from most subjects after tomato-soy cluster towards the left of the heatmap, where many features identified as soy isoflavone metabolites are present at higher abundances compared to the rest of the timepoints. When looking at the dendrogram from sample-based hierarchical clustering, the first branch represents post-tomato soy vs. the rest of the observations, suggesting this is the biggest differentiator in the data. Co-clustering of features that elicit similar responses is visualized, thus features that are more proximal are related in their metabolic profile. This finding reinforces the accurate annotations of soy isoflavones (as we would expect this group of compounds to generally act similarly) and provides context for potential relationships between unidentified features and parent/structurally similar compounds. Varying shades of red in each row of the heatmap for the soy isoflavone metabolites also illustrates interindividual variability of urinary isoflavone levels in subjects, as often observed in other human studies (Hutchins et al., 1995; Kelly et al., 1993; Lampe et al., 1998; Rowland et al., 2000). For isoflavones, the variability in responses is often hypothesized to be a result of differences in gut microflora (Frankenfeld et al., 2022; Mortensen et al., 2009; Soukup et al., 2023). For example, equol, a metabolite of daidzein, is produced by people with *Coriobacteriaceae* equol-producing bacteria (Clavel et al., 2014; Gong et al., 2023). Studies show that in Western populations, ∼25-30% of adults are “equol-producers” (Rowland et al., 2000; K. D. R. Setchell & Cole, 2006). In this study, we found 2 subjects (∼17%) with urine containing equol metabolites (**Figure S5, Supplementary Material**), similar to previous reports. To further illustrate individual heterogeneity upon tomato-soy consumption, prostate cancer patients who consumed the juice in a previous study were grouped into urinary daidzein metabolic phenotype clusters based on their proportions of urinary daidzein, dihydrodaidzein, O-DMA, and equol (Grainger et al., 2019). Out of the 33 subjects consuming tomato-soy juices, urinary equol was detected in 7 men (∼21%), who tended to produce a lower proportion of urinary O-DMA compared to subjects who do not produce equol (Grainger et al., 2019). In a cohort of men with prostate cancer fed soy-enriched bread, there were two equol-producing groups: one group produced proportionate amounts of equol and O-DMA, while the other produced very low levels of O-DMA (Ahn-Jarvis et al., 2015).

**Figure 6.**
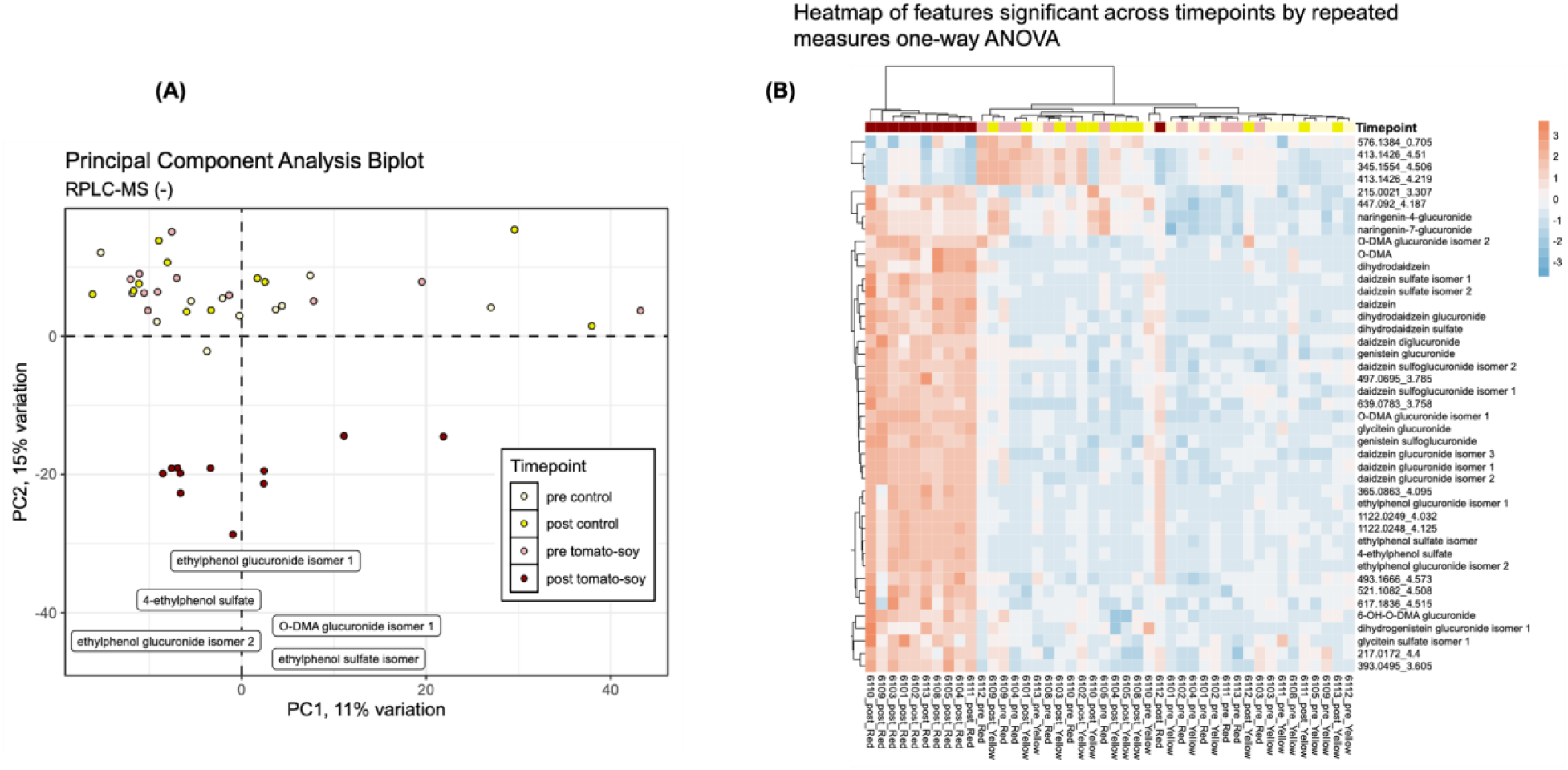
(A) Principal component analysis biplot for all intervention timepoints in each subject from RPLC-MS (-) untargeted metabolomics dataset (excluding quality control samples). (B) Heatmap of RPLC-MS (-) untargeted metabolomics data significant by repeated measures one-way ANOVA (FDR-corrected *P*-values < 0.05). Hierarchical clustering using Euclidean distances and Ward’s linkage method was used to categorize features vertically and samples horizontally.

Interestingly, 1 out of the 2 equol-producers in our study was the only subject to exhibit lower levels of O-DMA and glucuronide conjugate (**Figure S5, Supplementary Material**) following tomato-soy juice intake.

Among the key features altered by lycopene/soy intake in RPLC-MS (-) metabolomics analysis, almost half were identified as soy isoflavones with a level 2 or higher confidence (Figure 7, **Table 3** and **Table S2, Supplementary Material**) following an adapted version of the Metabolomics Standard Initiative guidelines (Sumner et al., 2007). Most flavonoids, including isoflavones, are more readily detected in LC-MS analysis using negative ionization mode when compared to positive mode (Cuyckens & Claeys, 2002, 2004; Rauha et al., 2001) due to the presence of hydroxyl groups which are easily ionizable. Soy isoflavones are excreted at high levels in the urine following intake, and their preferential detection in RPLC-MS (-) is consistent with their chemical properties and expected ionization behavior. We were though able to detect isoflavones metabolites as key features in the other three metabolomics datasets (see **Table S2, Supplementary Material**).

**Figure 7.**
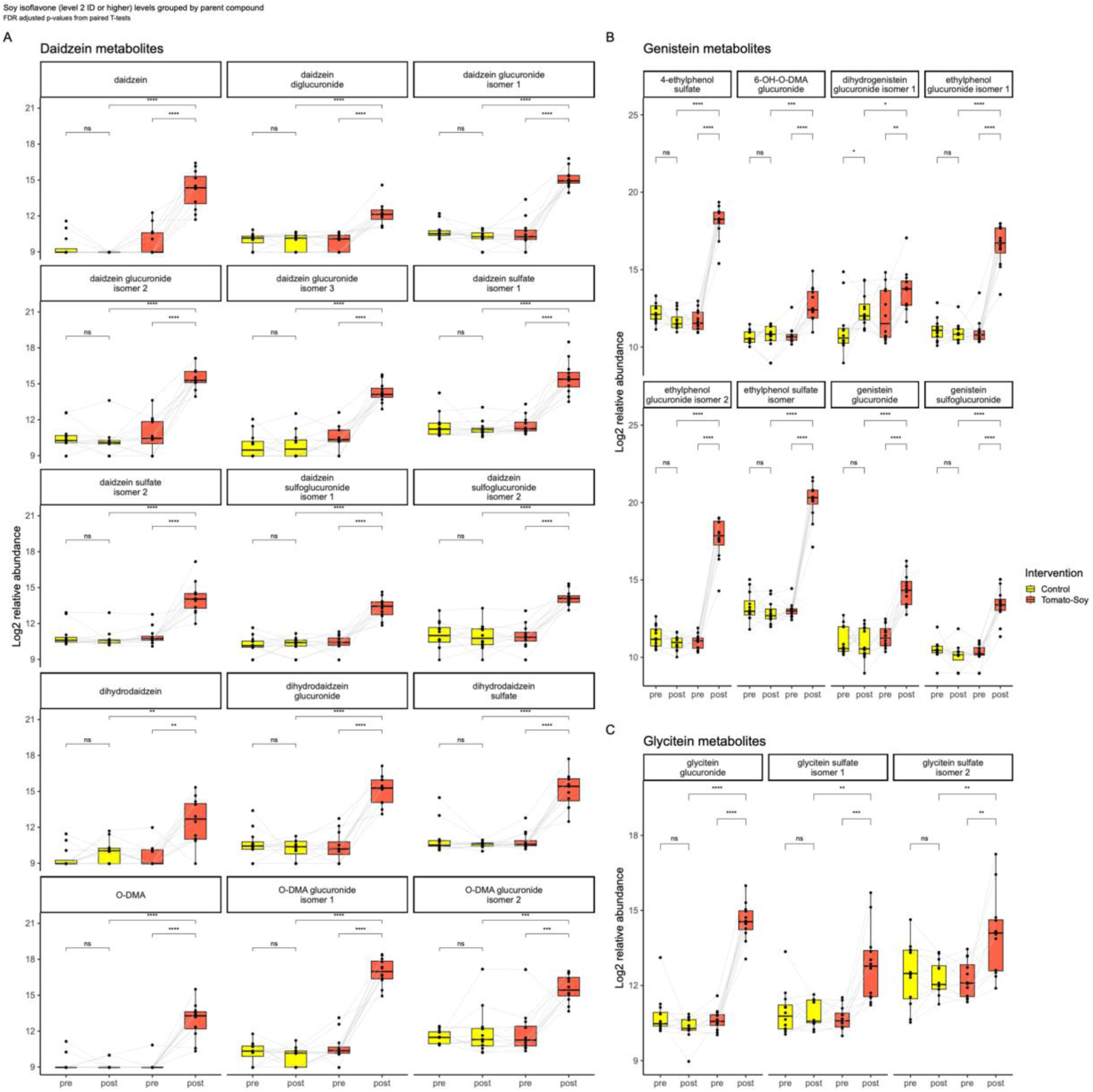
Boxplots of relative abundance levels for soy isoflavones and their metabolites detected in the urine, grouped by their parent compounds A) daidzein, B) genistein, and C) glycitein. *ns: not significant, *P < 0.05, ** P ≤ 0.01, *** P ≤ 0.001, **** P ≤ 0.0001*

**Table 3.** Isoflavone-derived metabolites that increased in urine (FDR-corrected *P* > 0.05, fold increase > 1.5) following tomato-soy intervention; detected in RPLC-MS (-) metabolomics dataset. *Feature identification levels are based on Metabolomics Standard Initiative guidelines (Sumner et al., 2007)*. *Features with ID level 1.5 were confirmed with biosynthesized standards*.

| Chemical name | Molecular formula | Retention time (min) | Theoretical $m/z$ [M-H] | Observed $m/z$ [M-H] | Mass error (Δppm) | Select MS/MS fragments | ID level | Additional isomers |
| --- | --- | --- | --- | --- | --- | --- | --- | --- |
| Daidzein | C<sub>15</sub>H<sub>10</sub>O<sub>4</sub> | 5.05 | 253.0506 | 253.0503 | 1.19 | 224.0476, 225.0563, 197.0605, 209.0607, 208.0531, 135.0070 | 1 | 0 |
| Daidzein glucuronide | C<sub>21</sub>H<sub>18</sub>O<sub>10</sub> | 4.03 | 429.0827 | 429.083 | 3.2 | 253.0507, 113.0245, 175.0252, 117.0187, 103.0039, 187.0049 | 1.5 | 2 |
| Daidzein diglucuronide | C<sub>27</sub>H<sub>26</sub>O<sub>16</sub> | 3.02 | 605.1148 | 605.1126 | 3.62 | 429.0820, 113.0243, 187.0061, 253.0502, 366.0352, 240.3893, 212.0010 | 2 | 0 |
| Daidzein sulfate | C<sub>15</sub>H<sub>10</sub>O<sub>7</sub>S | 4.33 | 333.0074 | 333.0047 | 8.17 | 253.0509, 254.0535, 315.1810 | 1.5 | 1 |
| Daidzein sulfoglucuronide | C<sub>21</sub>H<sub>18</sub>O<sub>13</sub>S | 0.7 | 509.0395 | 509.0384 | 2.19 | 333.0055, 253.0489, 429.0791, 113.0250, 175.0243, 312.9498 | 1.5 | 1 |
| Dihydrodaidzein | C<sub>15</sub>H<sub>12</sub>O<sub>4</sub> | 5.08 | 255.0663 | 255.0674 | -4.47 | 149.0246, 135.0088, 121.0294, 119.0502 | 1 | 0 |
| Dihydrodaidzein glucuronide | C<sub>21</sub>H<sub>20</sub>O<sub>10</sub> | 3.91 | 431.0983 | 431.0979 | 1.04 | 255.0656, 113.0243, 149.0244, 175.0247, 103.0035 | 1.5 | 0 |
| **Dihydrodaidzein sulfate** | C<sub>15</sub>H<sub>12</sub>O<sub>7</sub>S | 4.36 | 335.0231 | 335.0244 | -3.95 | 255.0664, 149.0246, 253.0508, 135.0086 | 1.5 | 0 |
| **O-DMA** | C<sub>15</sub>H<sub>14</sub>O<sub>4</sub> | 6.06 | 257.0819 | 257.0816 | 1.17 | 109.0296, 136.0169, 108.0216, 135.0088, 239.0716, 119.0502 | 1 | 0 |
| **O-DMA glucuronide** | C<sub>21</sub>H<sub>22</sub>O<sub>10</sub> | 4.89 | 433.114 | 433.113 | 2.31 | 257.1760, 113.0244, 415.1970, 127.0764, 117.0193 | 1.5 | 1 |
| **6-OH-O-DMA glucuronide** | C<sub>21</sub>H<sub>22</sub>O<sub>11</sub> | 4.3 | 449.1089 | 449.1074 | 3.37 | 273.0768, 113.0244, 351.1633, 175.0252, 193.0350, 431.1907 | 1.5 | 0 |
| **4-Ethylphenol sulfate** | C<sub>8</sub>H<sub>10</sub>O<sub>4</sub>S | 4.09 | 201.0227 | 201.0227 | -0.1 | 121.0657, 106.0422, 180.9892, 186.0004 | 1 | 1 |
| **Ethylphenol glucuronide** | C<sub>14</sub>H<sub>18</sub>O<sub>7</sub> | 4.5 | 297.098 | 297.0975 | 1.51 | 121.0659, 113.0244, 117.0192, 129.0193, 106.0423, 175.0252 | 1.5 | 1 |
| **Genistein glucuronide** | C<sub>21</sub>H<sub>18</sub>O<sub>11</sub> | 4.48 | 445.0776 | 445.0782 | -1.31 | 269.0458, 225.0562, 176.0120, 139.1143, 116.9941, 121.0659 | 1.5 | 0 |
| **Genistein sulfoglucuronide** | C<sub>21</sub>H<sub>18</sub>O<sub>14</sub>S | 3.61 | 525.0344 | 525.0334 | 1.97 | 349.0018, 269.0462, 445.0785, 113.0247, 175.0256 | 1.5 | 0 |
| **Dihydrogenistein glucuronide** | C<sub>21</sub>H<sub>20</sub>O<sub>11</sub> | 4.19 | 447.0933 | 447.092 | 2.83 | 271.0611, 113.0242, 175.0241, 151.0034, 129.0185, 433.5521 | 1.5 | 1 |
| **Glycitein glucuronide** | C<sub>22</sub>H<sub>20</sub>O<sub>11</sub> | 3.8 | 459.0933 | 459.093 | 0.58 | 283.0611, 113.0242, 187.0047, 193.0038, 175.0028, 131.0340, 441.1254 | 1.5 | 0 |
| **Glycitein sulfate** | C<sub>16</sub>H<sub>12</sub>O<sub>8</sub>S | 3.93 | 363.018 | 363.021 | -8.29 | 113.0240, 237.1132, 283.0614, 187.0981, 207.1028, 175.0246 | 1.5 | 1 |

Of the key features specific to lycopene/soy intake, we found that ethylphenol sulfate isomers resulted in the largest mean fold change amongst participants when comparing post-tomato-soy to both pre-tomato-soy (∼96-173-fold increase) and post-control (∼93-166-fold increase) (Figure 7B). 4*-*Ethylphenol was discovered as the major metabolite of both genistein and biochanin A in deconjugated sheep urine in the 1960s (Braden et al., 1967). 4-Ethylphenol glucuronide and sulfate conjugates have also been detected in rat urine following genistein administration (Paul et al., 2017; Yasuda et al., 2001). Not unique to isoflavone intake, ethylphenols have been reported to be produced from various phenolic compounds. In red wines, they are fermentation products formed in the presence of hydroxycinnamic acids such as *p*-coumaric acid and ferulic acid (Chatonnet et al., 1992, 1995). Additionally, urinary 4-ethylphenol significantly increased (1.1-fold increase) in healthy men following an oral dose of a flavonoid mixture (i.e., quercetin, epicatechin, and epigallocatechin gallate) (Loke et al., 2009). 4-Ethylphenol could be distributed peripherally following isoflavone intake, as it has been reported in rat tissues, including the brain following intraperitoneal genistein administration (Gamache et al., 1999; K. D. Setchell, 1998), and in plasma (Poulsen et al., 2009). To the best of our knowledge, we report intact ethylphenol conjugates (i.e., ethylphenol sulfates and glucuronides) in human urine following soy isoflavone intake for the first time. Ethylphenol compounds have also been reported to exert biological effects related to inflammation such as reduced fever and pain induced by lipopolysaccharide in rats (Yasuda et al., 2005) and stimulatory effects on prostaglandin F2α in vitro (Woclawek-Potocka et al., 2005). However, further research is needed to determine absolute levels and the clinical significance of ethylphenol conjugates in humans after soy consumption.

In this work, lycopene-derived metabolites were not found in urine. It is possible that the filtering criteria applied to the data, such as the fold change threshold, could have been too strict to capture low abundance metabolites, or our mass spectrometry methods were not sensitive enough. Lycopene metabolism is not fully understood, and there are no studies that have identified any specific metabolites in the urine. One small study administered a single dose of ^14^C-labeled lycopene to two men, and found radioactively detected metabolites in the urine, but these compounds were not identified (Ross et al., 2011). A very small percentage of total isotope was observed as unidentified polar products of lycopene in rat prostate following ^14^C-labeled lycopene supplementation (Zaripheh et al., 2003; Zaripheh & Erdman, 2005), suggesting that polar lycopene metabolites could be produced at low concentrations in mammalian tissue. There were unidentified metabolites in our work that were increased in response to tomato-soy intake that may be lycopene derivatives, though here we spent significant effort to do just this (e.g., selecting features significantly different between both pre- and post-tomato soy and post-control and post-tomato soy which had a fold change of 1.5 or greater, investigation of mass fragmentation spectra, comparing fragments to available spectra in the literature, and subjecting intervention juices to in vitro metabolism to produce corresponding features), without any success. The absence of detectable/identified lycopene metabolites in our method does not rule out their bioactivity in the tomato-soy intervention. Instead, it highlights both methodological limitations and gaps in current knowledge regarding the metabolic pathways of lycopene.

These findings suggest that the metabolomic response in urine to tomato-soy intake is driven in part or largely by soy-derived compounds, which may contribute to the observed anti-inflammatory changes.

#### 3.4.4 Independent of lycopene, tomato juice modestly alters the urine metabolome

Metabolites altered in the same direction following both tomato-soy and low-carotenoid tomato juice interventions suggests an effect of the tomato juice matrix independent of lycopene (and soy isoflavones). Represented in HILIC-MS (-) and RPLC-MS (+/-) datasets, naringenin glucuronides were significantly increased in the urine by ∼1.5 to 4-fold following both tomato interventions (**Table S3, Supplementary Material**), with control juice eliciting larger increases. A list of features annotated at a level 3 confidence or higher can be seen in **Table 4**. In RPLC-MS (-), naringenin-7-*O*-β-glucuronide was confirmed with authentic standard (level 1), while the other isomer was confirmed based on MS/MS spectral behavior (**Table 4**). Naringenin-related compounds (naringenin, chalconaringenin and respective glycosides) make up a large portion of flavonoids present in tomatoes (Slimestad et al., 2008; Slimestad & Verheul, 2009), making them plausible precursors for naringenin glucuronides detected in the urine here. Naringenin chalcone, an isomer of naringenin with an uncyclized C ring is reported to account for ∼65% of flavonoids quantified in fresh tomatoes (Slimestad et al., 2008), and has also been reported to produce naringenin glucuronides in rat urine following oral supplementation (Yoshimura et al., 2009). Additionally, naringin (a naringenin diglycoside) was found to undergo extensive metabolism after oral administration, yielding a number of conjugates including naringenin glucuronides in human urine (Zeng et al., 2017). While this is the first comprehensive report of the urine metabolome following tomato-based interventions in humans, targeted studies detect naringenin glucuronide as one of the main phenolics in blood and urine following tomato intervention (Martínez-Huélamo et al., 2016). Evidence also shows that following processed tomato juice consumption, naringenin glucuronide is more elevated in the plasma and urine of healthy subjects when compared to fresh tomato consumption (Martínez-Huélamo et al., 2016).

**Table 4.** List of features detected in ESI (-) that are significantly altered in the urine metabolome in the same direction following both low carotenoid yellow tomato juice and high lycopene tomato-soy juice consumption (P < 0.05, |fold change > 1.5|). Data was collected with two chromatography techniques: RPLC (reverse-phase liquid chromatography) and HILIC (hydrophilic interaction liquid chromatography). *Feature identification levels are based on Metabolomics Standard Initiative guidelines (Sumner et al., 2007). Features with ID level 1.5 were confirmed with biosynthesized standards*.

| Annotation | Molecular formula | Neutral mass (Da) | RPLC/HILIC | Retention time (min) | Theoretical $m/z$ [M-H] | Observed $m/z$ [M-H] | Mass error (Δppm) | Select MS/MS fragments | ID level |
| --- | --- | --- | --- | --- | --- | --- | --- | --- | --- |
| ***Flavonoid metabolites*** |  |  |  |  |  |  |  |  |  |
| Naringenin-7-*O*-β-glucuronide | C<sub>12</sub>H<sub>20</sub>O<sub>11</sub> | 448.1006 | RPLC | 4.50 | 447.0933 | 447.0919 | 3.05 | 271.0608, 113.0241, 175.0245, 151.0034, 201.0220, 429.1365, 351.1613 | 1 |
| Naringenin glucuronide isomer | C<sub>12</sub>H<sub>20</sub>O<sub>11</sub> | 448.1006 | RPLC | 4.07 | 447.0933 | 447.0921 | 2.60 | 271.0611, 113.0239, 273.0768, 187.0067, 175.0246, 272.0656, 165.0197, 237.0078, 429.1361 | 2 |
| ***Phenolic acid catabolites and their conjugates*** |  |  |  |  |  |  |  |  |  |
| Acetylphenol sulfate | C<sub>8</sub>H<sub>8</sub>O<sub>5</sub>S | 216.0092 | HILIC | 1.17 | 215.0019 | 215.0018 | 0.68 | 135.0448, 126.9950, 129.0359, 201.0236 | 3 |
| Dihydroxybenzoic acid | C<sub>7</sub>H<sub>6</sub>O<sub>4</sub> | 154.0266 | HILIC | 3.42 | 153.0193 | 153.0193 | 0.07 | 108.0220, 109.7921, 112.7246, 128.0510, 137.2723 | 2 |
| Vanillin sulfate | C<sub>8</sub>H<sub>8</sub>O<sub>6</sub>S | 232.0042 | HILIC | 1.61 | 230.9969 | 230.9967 | 0.70 | 151.0402, 109.0490, 136.0529, 129.0343 | 2 |
| Dihydrocaffeic<br>acid sulfate or<br>homovanillic acid<br>sulfate | $C_9H_{10}O_7S$ | 262.0147 | HILIC | 0.74 | 261.0074 | 261.0072 | 0.87 | 166.0268, 181.0502,<br>179.0342, 138.0329,<br>125.0611, 216.7764,<br>151.0391 | 2 |
| Tyrosol<br>glucuronide | $C_{14}H_{18}O_8$ | 314.1002 | HILIC | 5.70 | 313.0929 | 313.0919 | 3.10 | 137.0604, 176.9846,<br>258.9239, 135.0298,<br>276.8748, 119.0489 | 1.5 |

Free naringenin and its conjugates including glucuronides have been found in rat tissues such as the liver, kidneys, spleen, heart, adipose and lungs (El Mohsen et al., 2004; Lin et al., 2014; Zou et al., 2012). These findings inform that naringenin-derived compounds are absorbed and may be transported to tissues, thus having the ability to exert localized biological effects. In a feeding study, naringenin-supplemented diets were shown to exert anti-inflammatory effects in adipose tissue of high-fat diet-fed mice by significantly inhibiting toll-like receptor 2 (TLR2), TNF-α, and MCP-1 expression (Yoshida et al., 2013). Additionally, a cell study found that physiological levels (∼600 nmol/L) of naringenin and its glucuronides (naringenin-7-*O*-glucuronide and naringenin-4ʹ-*O*-glucuronide) perturbed inflammatory human macrophage gene expression (Dall’Asta et al., 2013). The increase of naringenin glucuronide levels in urine following both tomato interventions observed in this study could suggest that these compounds contribute to the overall biological effects of the tomato-based interventions used in our work.

Outside of naringenin glucuronide isomers, we did not find flavonoid aglycones and other flavonoid conjugates as key features associated with tomato-based intake. Rutin (quercetin-3-rutinoside) is another major flavonoid present in tomatoes (Slimestad et al., 2008), although abundant in the peel with trace amounts in the flesh (Bovy et al., 2007). Quercetin can arise from the enzymatic/microbial cleavage of the rutinoside sugar in the large intestine. However, quercetin and its conjugates did not emerge as significant features following tomato-based interventions in our work. This aligns with previous studies that have reported trace levels of quercetin in the urine (Martínez-Huélamo et al., 2016), suggesting limited absorption or further microbial metabolism into smaller compounds such as phenolic acids (Jaganath et al., 2006; Mullen et al., 2008). Furthermore, during tomato processing the peel is removed and thus low levels of rutin would be expected in the juices, and consequentially lower levels of rutin-derived metabolites in the body following consumption. Hydroxycinnamic acids such as chlorogenic acid, caffeic acid, and their derivatives (e.g., sugars, quinic acid conjugates) are also major polyphenols found in tomato (Gómez-Romero et al., 2010; J. Li et al., 2024). These compounds undergo substantial metabolism by intestinal microbiota as well, yielding a diversity of phenolic catabolites (Hasyima Omar et al., 2021; Morishita & Ohnishi, 2001). In this study, several features were putatively annotated as phenolic catabolites (and their conjugates): acetylphenol sulfate, protocatechuic acid, vanillin sulfate, homovanillic acid sulfate, and tyrosol glucuronide, which were significantly elevated following both tomato-based interventions (**Table 4, Table S3, Supplementary Material**). Additionally, three features associated with tomato intake (*m/z* 234.1122, 250.1107, and 292.1575) were detected in RPLC-MS (+) (**Table 5**). Based on the presence of characteristic fragments corresponding to a cinnamoyl moiety (*m/z* 131 and 103), we hypothesize these to be cinnamic acid derivatives (Level 3 annotation) (**Table 5**, **Figure S6, Supplementary Material**). Fragments 131 and 103 have been reported for compounds with a cinnamic acid backbone such as hydroxycinnamic acids, caffeoyl alkaloids, and iridoid glycosides (P.-Y. Chen et al., 2016; Du et al., 2019; Es-Safi et al., 2007; Z. Li et al., 2018; Zhang et al., 2015). While the MS/MS spectral properties for *m/z* 250 differed most from the other two features (*m/z* 234 and 292), this mass was present in the fragmentation spectra for *m/z* 292 (see **Table 5**), increasing plausibility that *m/z* 250 is structurally similar to this triad of features. We suspect that these compounds are amide conjugates because of the difference between *m/z* 188.1070 and 131.0493 observed in the MS/MS spectra for *m/z* 234 being 57.0577, which corresponds to protonated allylamine (theoretical *m/z* 57.0578) and thus could suggest the presence of an amine-containing group (**Figure S6, Supplementary Material**). Additionally, the formulas calculated using ChemCalc (Patiny & Borel, 2013) were all within ∼10 ppm for each feature, and all included one nitrogen. The three metabolites putatively annotated as cinnamic acid amides 1-3 (*m/z* 234.1122, 250.1107, and 292.1575, respectively) (Figure 8) seem to be related due to their spectral similarities (see **Table 5**) and co-clustering (**Figure S7, Supplementary Material**). *m/z* 250 is hypothesized to be the hydroxylated (+16 *m/z*) structure of *m/z* 234, while we suspect *m/z* 292 to be an acetylated form of *m/z* 250 (+42 *m/z*). These three metabolites significantly increased (Figure 8) ranging from ∼4-10 fold-change increase from pre- to post-tomato intervention, with the control intervention inducing a relatively higher fold increase (Figure 8). In the liver and kidneys, glycine-N-acyltransferase (GLYAT), mediates the conjugation of glycine to facilitate the removal of aromatic acids–many of which are produced by gut microbial-derived dietary polyphenols (Badenhorst et al., 2014; Beyoğlu & Idle, 2012; Leung et al., 2020). There are numerous reports of cinnamic acids being metabolized into hippuric acid (*N*-benzoylglycine) (Hoskins et al., 1984; Nutley et al., 1994; Snapper et al., 1940). Notably, in a discovery urine biomarker study, hippuric acids were amongst the highly ranked signals for habitual tomato intake (Beckmann et al., 2013). Other cinnamic acid-glycine conjugates such as feruloylglycine have been detected in human urine following coffee intake, and in rat urine (along with isoferuloylglycine) after caffeic acid consumption (Shi et al., 2019; Stalmach et al., 2009). A urine metabolomics study feeding humans cocoa powder also putatively identified vanilloylglycine as a discriminating feature following consumption (Llorach-Asunción et al., 2010). Glycine conjugates of cyclohexanecarboxylic acid and cyclohexadienecarboxylic acid contributed to the differences in urinary profiles following plant-rich diets low in flavonoids (Ulaszewska et al., 2016). The authors suggested these compounds to be intermediate products of cinnamate metabolism (Ulaszewska et al., 2016). We were unable to annotate these putative cinnamic acid amides here as glycine-conjugates. While glycine conjugates remain possible structures for these features, another plausible explanation is that gut microbes may transform glycine-conjugated cinnamic acids into unknown modified amine-containing derivatives. This is supported by evidence that gut microbes can modify aromatic amines through enzymes such as aromatic amino acid decarboxylase (Sugiyama et al., 2022). Hydroxycinnamic acid amides such as feruloyltyramine and caffeoylputrescine have been detected in tomato juices (Cichon, Riedl, & Schwartz, 2017), and could also be precursors to the features we annotate as cinnamic acid amides in this study. Additionally, out of all phenolic compounds quantified in tomato products, it was reported that phenolic acid conjugates (e.g., chlorogenic acid, ferulic acid hexoside, homovanillic acid hexoside) made up the largest proportion (Martínez-Huélamo et al., 2016).

**Figure 8.**
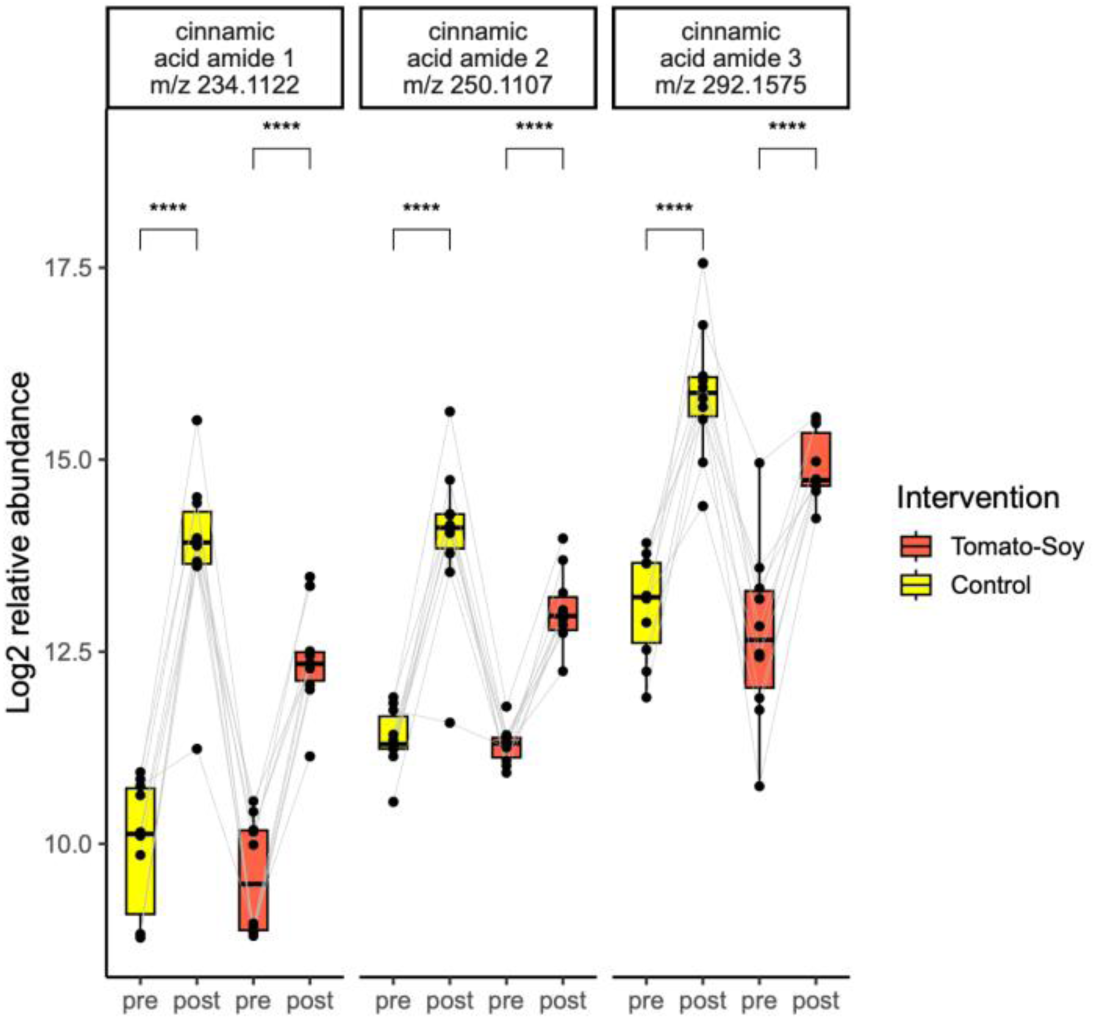
Boxplots of putative cinnamic acid derivatives in urine detected in RPLC-MS (+) dataset that increase significantly following both tomato-soy and control tomato juices. *\*\*\*\* P ≤ 0.0001*

**Table 5.** List of features detected in ESI (+) that are significantly altered in the urine metabolome in the same direction following both low carotenoid yellow tomato juice and high lycopene tomato-soy juice consumption (P < 0.05, |fold change > 1.5|). Data was collected with two chromatography techniques: RPLC (reverse-phase liquid chromatography) and HILIC (hydrophilic interaction liquid chromatography). *Feature identification levels are based on Metabolomics Standard Initiative guidelines (Sumner et al., 2007)*.

| Annotation | Molecular formula | Neutral mass (Da) | RPLC/<br>HILIC | Retention time (min) | Theoretical $m/z$ [M+H] | Observed $m/z$ [M+H] | Mass error (Δppm) | Select MS/MS fragments | ID level |
| --- | --- | --- | --- | --- | --- | --- | --- | --- | --- |
| *Phenolic acid catabolites* |  |  |  |  |  |  |  |  |  |
| Cinnamic acid amide 1 | C<sub>13</sub>H<sub>15</sub>NO<sub>3</sub> | 233.1052 | RPLC | 4.88 | 234.1125 | 234.1122 | 1.25 | 131.0495, 103.0541, 106.0142, 150.1289, 146.9905, 207.0611, 216.1124, 121.0991, 175.1122, 103.0623 | 3 |
| Cinnamic acid amide 2 | C<sub>13</sub>H<sub>15</sub>NO<sub>4</sub> | 249.1001 | RPLC | 3.55 | 250.1074 | 250.1107 | -13.16 | 122.0268, 130.0498, 145.0680, 158.1165, 104.9997, 191.0737 | 3 |
| Cinnamic acid amide 3 | C<sub>16</sub>H<sub>21</sub>NO<sub>4</sub> | 291.1471 | RPLC | 3.66 | 292.1544 | 292.1575 | -10.75 | 103.0577, 131.0534, 144.1031, 193.1220, 244.1530, 121.1031, 250.1112*, 175.1133 | 3 |
| *Endogenous metabolites* |  |  |  |  |  |  |  |  |  |
| Medium chain acylcarnitine (CAR 4:1;O2) | C<sub>11</sub>H<sub>19</sub>NO<sub>6</sub> | 261.1212 | HILIC | 5.30 | 262.1285 | 262.1284 | 0.53 | 262.1234, 203.0511, 171.5822, 144.1021, 103.0383 | 3 |

Altogether, the evidence discussed supports the plausibility that the tomato juices provided in this study provided phenolic precursor compounds–particularly cinnamic acid derivatives–that could give rise to the observed urinary features hypothesized to be cinnamic acid amides following host and microbial metabolism.

Beyond polyphenols, other tomato-derived compounds may also influence the efficacy of the tomato-based juices provided in this study. Alkaloids, particularly steroidal alkaloids and imidazole alkaloids are present in tomatoes (Dzakovich et al., 2020, 2022; Hövelmann et al., 2019) and have been found in mammalian biospecimens following consumption. Tomato steroidal alkaloids (e.g., alpha-tomatine, tomatidine, esculeogenin, and their metabolites) have been reported in human urine and plasma (Hövelmann, Jagels, et al., 2020; Hövelmann, Lewin, et al., 2020), pig plasma (Sholola et al., 2025), in addition to murine plasma, liver and skin (Cichon, Riedl, Wan, et al., 2017; Cooperstone et al., 2017; Dzakovich et al., 2024). Imidazole alkaloids, such as *N*-caprylhistidinol and its glucuronide conjugate, have also been detected in human urine following tomato consumption (Hövelmann, Lewin, et al., 2020). Neither steroidal nor imidazole alkaloids were selected as significant metabolites in the urine following tomato intake in our study. After querying known tomato alkaloid metabolite masses from extracted ion chromatograms, two peaks corresponding to steroidal alkaloid masses were detected in RPLC-MS (+) mode: *m/z* 416.3517 and 432.3465 (**Table S6, Supplementary Material**). These ions are accurate mass matches within 3 ppm to tomatidine and hydroxytomatidine, respectively. These features were significantly increased by ∼2-fold (*P* < 0.05) following the tomato-soy intervention when compared to before. Neither of these features were significantly changed following the control tomato intervention, which is why they were not selected when identifying important features related to tomato intake. After this finding, authentic tomatidine standard was analyzed alongside urine samples to confirm *m/z* 416, but the retention time was not a match. MS/MS spectra for both *m/z* 416 and 432 were investigated and did not exhibit characteristic steroidal alkaloid fragments reported in the literature (Caprioli et al., 2014, 2015; Cichon, Riedl, Wan, et al., 2017; Cooperstone et al., 2017; Dzakovich et al., 2020, 2024; Hövelmann, Jagels, et al., 2020; Hövelmann, Lewin, et al., 2020; Sholola et al., 2025) and thus we were unable to conclude the annotation of these features. The urine collected from subjects in this study were 24-hour urine collections, and were diluted for metabolite normalization and extraction. This could have resulted in alkaloid concentrations too low for detection with our method. Therefore, it shouldn’t be implied that alkaloids derived from tomatoes are absent in the urine following these interventions.

While lycopene and soy isoflavones represent the key bioactive compounds of interest from our tomato-soy juice product, we acknowledge that other constituents within the tomato juice matrix could contribute to metabolic and inflammatory changes post-consumption. For instance, both tomato interventions resulted in a significant 50% reduction of a putatively annotated medium chain acylcarnitine (**Table 5**) in the urine. Though we were unable to resolve specifically which medium chain acylcarnitine was decreased, the literature suggests that this group of fatty acid metabolites are often elevated in low-grade chronic inflammation (e.g., obesity and diabetes) (Adams et al., 2009; Mihalik et al., 2010; Rutkowsky et al., 2014). This suggests that phytochemicals independent of lycopene in tomato could be contributing to the anti-inflammatory effects of tomato-soy juice.

Investigating the health benefits of whole-food products is inherently complex due to the multitude of bioactive compounds present (and their potential interactions). However, this study design comparing high-lycopene tomato-soy juice to a control low-carotenoid tomato juice provides us the opportunity to disentangle the effects of lycopene and isoflavones from other components in the provided food matrix. By using a tomato-based control food, this approach is intended for a more granular investigation into the contributions of specific phytochemicals while still capturing the broader impact of dietary patterns. Importantly, the metabolites detected in the metabolomics portion of this work were not quantified, which must occur to understand their physiological relevance in this context. Still, we report metabolites that are significantly altered upon tomato juice consumption as a start.

## 4 Conclusion

This study demonstrates that consumption of tomato-soy juice, a tomato-based functional beverage enriched in lycopene and soy isoflavones, modulate systemic inflammatory markers and the urine metabolome in individuals with obesity after 4 weeks of daily consumption. We observed significant reductions in systemic pro-inflammatory cytokines: IL-12p70, IL-5, GM-CSF, as well as a trend towards reduced TNF-α. Circulating lycopene levels increased following the tomato-soy intervention to 1,298.4 nmol/L, significantly exceeding both pre-tomato-soy and post-control timepoints. Additionally, the urinary metabolome after tomato-soy intake was distinct from that of the control intervention, and this was largely due to increased excretion of soy isoflavones and their microbial metabolites, including ethylphenol sulfates derived from genistein and O-DMA glucuronides from daidzein. To the best of our knowledge, this is the first report of intact ethylphenol sulfates and glucuronides in the urine following soy intake. In this study, we also report the urine metabolome following a controlled tomato-based intervention for the first time.

The design of this trial was intended to capture the isolated effects of lycopene and soy isoflavones from the tomato juice matrix. While these compounds were significantly elevated post-tomato-soy, the reductions in pro-inflammatory cytokines were not different between post-tomato-soy and post-control timepoints. We speculate that this could be attributed to a lower-than-expected sample size, limiting statistical power. Additionally, the anti-inflammatory effects observed may not be solely attributable to lycopene and/or soy isoflavones as other phytochemicals present in tomato juice could contribute. For instance, both tomato interventions (tomato-soy and low-carotenoid control juices) led to a significant increase in the urinary excretion of naringenin glucuronides, phenolic acids, and their catabolites, as well as a significant reduction of a medium-chain acylcarnitine–a class of fatty acid catabolites linked to inflammatory processes which could underlie some of the effects seen. These findings underscore the benefit of complementary metabolomics analysis to uncover unanticipated contributors to complex dietary interventions.

By leveraging a crossover design and a whole-food approach, our study aimed to delineate the effects of lycopene and soy isoflavones in a tomato juice product that may not be achieved solely with isolated compounds. Originally developed for its beneficial properties in the context of prostate cancer (Grainger et al., 2008, 2019; Zuniga et al., 2013), our findings along with a preclinical chronic pancreatitis study (Mukherjee et al., 2020) support the broader potential of tomato-soy juice to influence immune-metabolic pathways relevant to chronic diseases. Future clinical and mechanistic studies will characterize the inflammatory and metabolic mechanisms of tomato-soy interventions.

## Data Availability

Raw MS1 data are available in Metabolomics Workbench (ST004178) and data-dependent MS/MS data have been reposited with GNPS MassIVE (MSV000099128). Code for metabolomics, carotenoids, and cytokines analyses can be found at https://github.com/CooperstoneLab/TomatoSoy-Obesity-Metabolomics.

## Acknowledgements

This work was supported by grants from the United States Department of Agriculture (NIFA AFRI 2018-67017-27519, National Needs Fellowship 2020-38420-30723, and Hatch OHO01470, OHO01563, and OHO01538) and the National Institutes of Health (R01DK138871). Research was additionally supported by Lisa and Dan Wampler Endowed Fellowship for Foods and Health Research, and the Foods for Health Initiative at the Ohio State University.

